# The efficacy of coenzyme Q_10_ treatment in alleviating the symptoms of primary coenzyme Q_10_ deficiency: a systematic review

**DOI:** 10.1101/2022.05.21.22275418

**Authors:** Ying Wang, Siegfried Hekimi

**Affiliations:** Department of Biology, McGill University, Montreal, Quebec, Canada

**Keywords:** Coenzyme Q, ubiquinone, CoQ_10_ supplementation, primary CoQ_10_ deficiency, CoQ biosynthesis, mitochondrial disorder

## Abstract

Coenzyme Q_10_ (CoQ_10_) is necessary for mitochondrial electron transport. Mutations in CoQ_10_ biosynthetic genes cause primary CoQ_10_ deficiency (PCoQD) and manifest as mitochondrial disorders. It is often stated that PCoQD patients can be treated by oral CoQ_10_ supplementation. To test this, we compiled all studies describing PCoQD patients up to May 2022. We excluded studies with no data on CoQ_10_ treatment, or with insufficient description of effectiveness. Out of 303 PCoQD patients identified, we retained 89 cases, of which 24 reported improvements after CoQ_10_ treatment (27.0%). In five cases, the patient’s condition was reported to deteriorate after halting of CoQ_10_ treatment. 12 cases reported improvement in the severity of ataxia, and 5 cases in the severity of proteinuria. Only a subjective description of improvement was reported for four patients described as responding. All reported responses were partial improvements of only some symptoms. For PCoQD patients, CoQ_10_ supplementation is replacement therapy. Yet, there is only very weak evidence for the efficacy of the treatment. Our findings thus suggest a need for caution when seeking to justify the widespread use of CoQ_10_ for the treatment of any disease or as dietary supplement.

**Highlights:**

1. Only 27% of primary CoQ_10_ deficiency patients benefited from CoQ_10_ supplementation.
2. Studies of the effects of supplementation necessarily lacked controls and blinding.
3. All reported positive responses to treatment only partially improved few symptoms.
4. CoQ_10_ supplementation for the treatment of any disease should be questioned.
5. Firm evidence of benefits requires randomize, controlled trials of CoQ_10_ therapy.

**Graphic Abstract:** 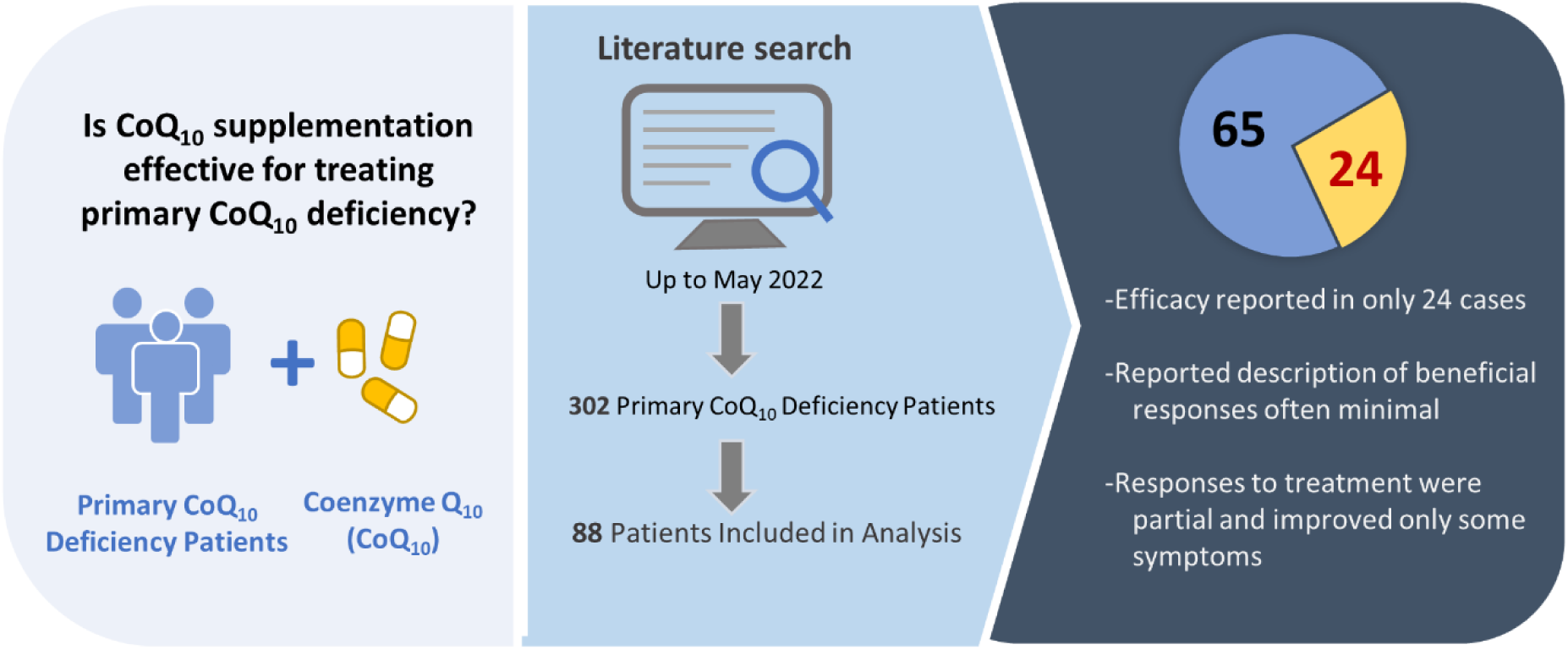

## 1. Introduction

Coenzyme Q_10_ (CoQ_10_), also known as ubiquinone (UQ_10_), is composed of a redox active aromatic ring and a ten-repeat long polyprenyl sidechain. CoQ_10_ is an essential component of the mitochondrial respiratory chain, where it functions as a mobile carrier for the transfer of electrons from respiratory complexes I and II to complex III, and as cofactor in complex III function. In addition, it feeds electrons into the respiratory chain from other entry points, including the electron transfer flavoprotein, sulfide-quinone reductase, and dihydroorotate dehydrogenase [1-4]. CoQ_10_ is known to have antioxidant properties and to be involved in several other cellular functions outside of mitochondria [5, 6]. As far as is known, all cells rely exclusively on endogenous CoQ synthesis. So far, 11*COQ* genes whose products participate in CoQ_10_ biosynthesis have been identified in humans. Some of them function as enzymes and others as structural components of the CoQ biosynthetic complex (Fig. 1A) [7-11]. Mutations in *COQ* genes cause primary CoQ_10_ deficiency (PCoQD), a clinically heterogenous and rare disorder [12, 13]. Symptoms often resemble those of typical inborn mitochondrial respiratory chain disorders (Fig. 1B), including early onset, multi-organ involvement, and with prevalent neurological and muscular manifestations. It some cases the symptoms predominantly affect a particular organ or tissue (e.g., kidney- or cerebellum-limited phenotypes) [8, 14-16]. Secondary CoQ_10_ deficiency refers to all the conditions in which the etiology of a CoQ_10_ deficiency is not a molecular lesion in the CoQ_10_ biosynthetic pathway [17, 18]. In fact, a variety of conditions have been found to be associated with CoQ_10_ deficiency. Statins were shown to reduce serum and muscle CoQ_10_ levels [19, 20]. Mutations in the electron transfer flavoprotein dehydrogenase (*ETFDH*), and mitochondrial DNA (mtDNA) lesions, including low mtDNA copy number, were also shown to lower steady state level of CoQ_10_ [21-26]. The mechanisms leading to deficiency in these cases are unknown, except for the effect of statins, which inhibit the synthesis of mevalonate, the molecular precursors of the CoQ_10_ sidechain.

**Figure 1.**
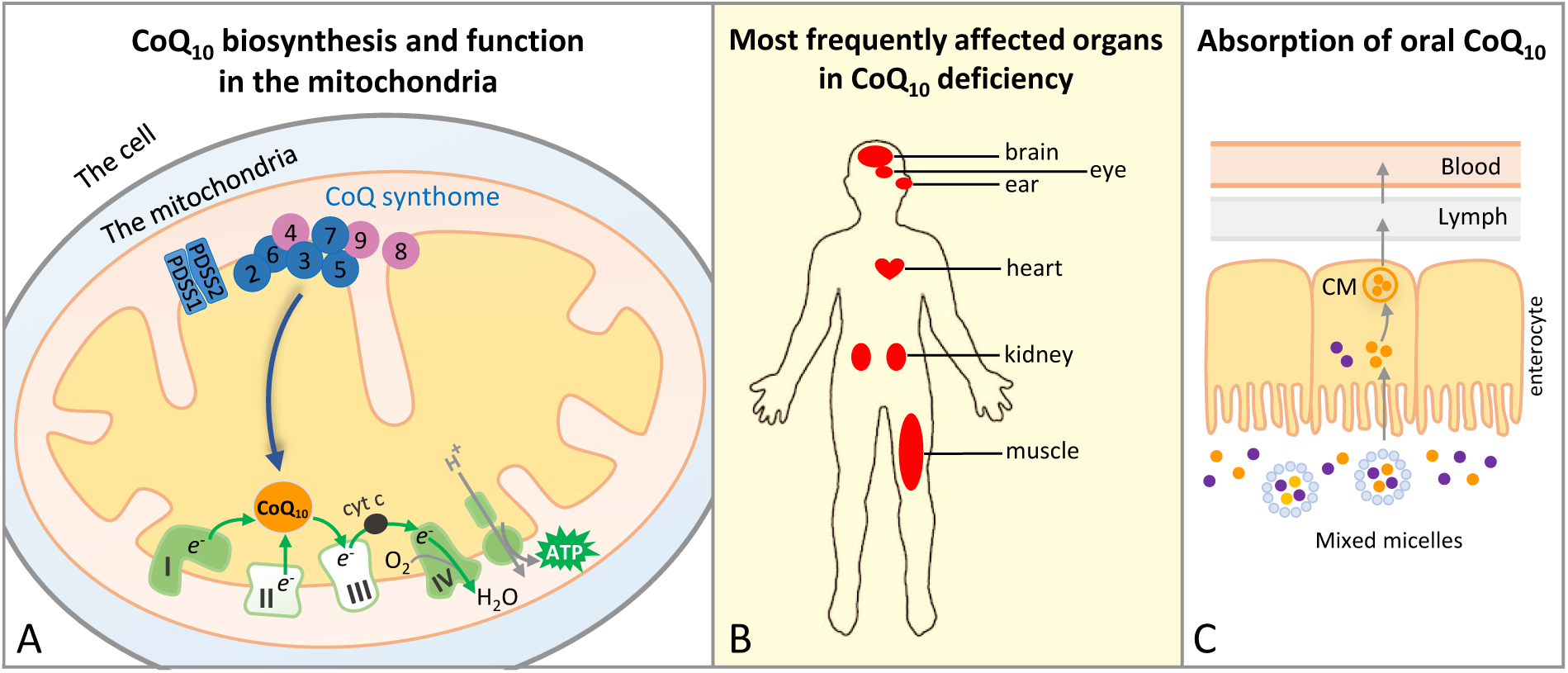
CoQ_10_ in the mitochondria, pathology of CoQ_10_ deficiency and oral supplementation. **(A)** The final steps of CoQ_10_ biosynthesis are carried out in the inner mitochondrial membrane. The CoQ_10_ biosynthetic pathway includes both enzymes (in blue) and structural or regulatory components (in purple). Only the numbers in their names are shown for COQ proteins (COQ2-7, COQ8A, COQ8B and COQ9). They are known to form a large complex, the CoQ biosynthetic complex or CoQ-synthome. COQ10A and COQ10B whose functions are uncertain and not known to be part of the complex are not shown. The most essential function of CoQ_10_ is to transport electrons in the mitochondrial respiratory chain. Although CoQ_10_ is found in the mitochondrial membrane, in the figure this is not shown for clarity. **(B)** Primary CoQ_10_ deficiency predominantly manifests as mitochondrial disorder, with organs with high energy needs being most often affected. **(C)** Intestinal absorption of CoQ_10_ is thought to occur through the formation of mixed micelles with other dietary lipids. Once inside the enterocytes, CoQ_10_ is incorporated into chylomicrons (CM) which are transported via the lymphatics to the blood circulation. Because of its extreme hydrophobicity and its relatively large size, the absorption of orally administered CoQ_10_ has been reported to be poor.

In tissue samples or cultured cells from patients, CoQ_10_ deficiency can be diagnosed by measuring CoQ_10_ levels, which can be complemented by the observation of impaired CoQ_10_-dependent respiratory chain activities (Complex I-III and Complex II-III). In the last few decades, with the increasing availability and affordability of genomic sequencing technology, whole genome or exome sequencing is increasingly becoming the first-line diagnostic test for patients suspected of having genetic disorders, including PCoQD. This has accelerated the discovery of novel PCoQD disease variants [27]. Disease-causing mutations have been reported for 9 out of 11 *COQ* genes required for CoQ biosynthesis. Below we report that at least 303 PCoQD patients have been reported so far. CoQ_10_ supplementation is frequently initiated immediately after diagnosis (Fig. 1C), and the majority of the literature on CoQ_10_ deficiency states that CoQ_10_ deficiency is treatable by supplementation with exogenous CoQ_10_ [28-33]. However, there is lack of clear evidence for such a claim.

In addition to patients with documented CoQ_10_ deficiency and/or *COQ* mutations, CoQ_10_ is frequently recommended to mitochondrial disease patients [34, 35]. In fact, it is a component of the so-called mitochondrial cocktail which is a collection of high-dose nutraceuticals with the potential to support mitochondrial functioning [36]. Moreover, CoQ_10_ is recommended for treating a wide range of other conditions (e.g., heart failure and neurodegenerative diseases) and it is widely available over the counter as an anti-aging dietary supplement [32]. By estimation, the global market size of CoQ_10_ amounts to close to 600M USD a year.

This review aims to summarize and evaluate the available evidence for the effectiveness of CoQ_10_ supplementation for the treatment of PCoQD. Patients with PCoQD should be the most amenable to CoQ_10_ treatment because their CoQ_10_ deficiency is the only cause of all their symptoms and therefore CoQ_10_ treatment is simple replacement therapy. Thus, examining outcomes of CoQ_10_ treatment for these patients is the first key step to address the effectiveness of any CoQ_10_ therapy and to promote a rational use of CoQ_10_ for disease treatment or as a health supplement.

## MATERIALS AND METHODS

### Search strategy and selection criteria

A literature search was performed in PubMed for studies that described PCoQD patients, up until May 01, 2022. The PubMed query used is given in Supporting Information. The references cited in the articles identified were manually screened for any additional relevant study. We imposed no publication status or language restrictions. We considered any type of study regardless of research design.

The following information was sought in each paper: descriptive characteristics of PCoQD patients including sex, age of onset, major symptoms, age at the last reported exam or death, molecular lesions in *COQ* genes or proteins, severity of CoQ_10_ deficit, respiratory chain complex (RCC) activities, CoQ_10_ treatment received and clinical outcomes, and laboratory tests known to be relevant to mitochondrial disease. CoQ_10_ levels and RCC activities are most often reported in patient-derived skin fibroblasts or muscle biopsies. Study data were extracted by one reviewer (YW) and verified by another reviewer (SH) for accuracy, narrative summaries, and interpretation. When data were reported more than once for the same patients, which was exceedingly rare, the data that were included were those from the most recent comprehensive report. If no data on patient treatment with CoQ_10_ was provided in a study, or if patients were treated but outcome data was not reported, or the reported effects were contradictory or ambiguous, the study was excluded from the final data synthesis (Fig. 2).

**Figure 2.**
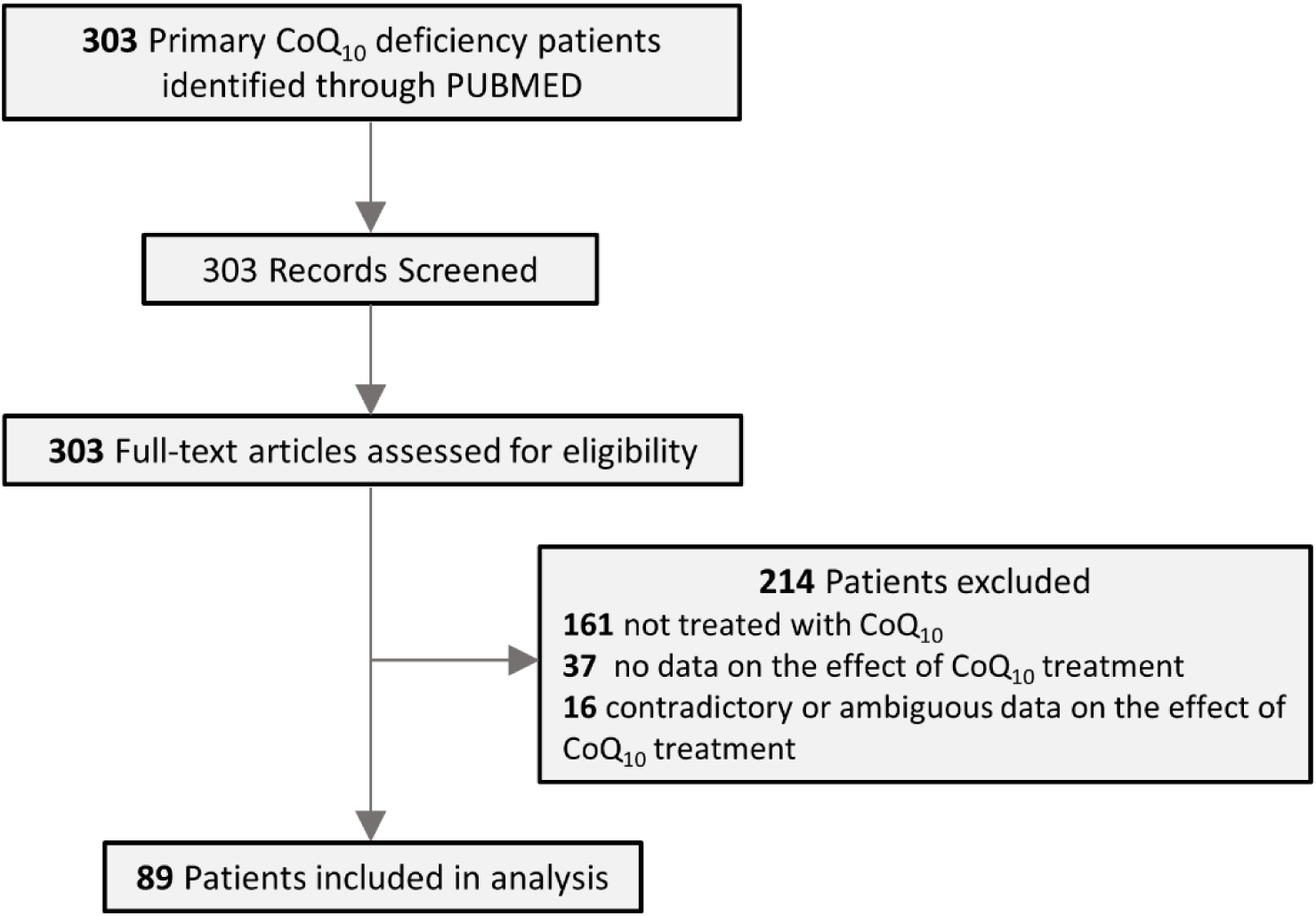
Flow diagram for identification and selection of primary CoQ_10_ deficiency patients.

### Data analysis

We synthesized data using tabulations that include narrative summaries. The effect of CoQ_10_ treatment on clinical outcomes is considered as positive (responding) if one of the following criteria is satisfied: a) a positive effect on a quantifiable clinical measure was reported; b) some improvement was noted after CoQ_10_ treatment and stopping/halting the treatment resulted in deterioration of a patient’s condition; and/or c) no quantifiable clinical evidence was provided but at least two symptoms were described to be improved following CoQ_10_ treatment. Fulfilling any one of the first two criteria is defined as responding with an objective description of the response. Whereas if symptom improvement was described without relying on any quantifiable measure, we categorize it as a subjective description of the response to CoQ_10_ therapy. Patients counted as not-responding include cases where no significant effect was noted after CoQ_10_ treatment, or the reported effect(s) were minimal, or when, though some clinical improvement was noted, the patient’s condition had deteriorated (e.g., developed new symptoms) while on CoQ_10_ therapy. No restriction on CoQ_10_ dosage (dose, formulation, dose frequency), time of initial treatment, duration of treatment, or concurrent treatments was made. The two authors independently assigned the patient cases to the categories. Disagreements were resolved by discussion and consensus.

## RESULTS

The literature search yielded 78 published studies, from which a total of 303 patients with PCoQD were identified. Their characteristics are summarized in Table 1, and details are available in Table S1. Of the 303 PCoQD patients, 142 [46.7%] were reported to receive oral supplement of CoQ_10_. The dosage was reported as mg/day in some studies and as mg/kg/day in others. Doses ranged from 60 mg/day to 2,100 mg/day or from 5 mg/kg/day to 100 mg/kg/day, and the reported duration of treatment was from 1 month to 8 years [37-40]. Following the exclusion criteria, 53 treated patients were removed from the final analysis (Table S2). Among the excluded patients, 16 were excluded because the reported follow-up findings were judged to be ambiguous or inadequate to judge treatment efficiency, for example reports that mention symptom stabilization or CoQ_10_ treatment combined with other simultaneous treatments. All other exclusions were because no treatment outcome was reported.

**Table 1.** Primary CoQ_10_ deficiency patients reported in the literature.

| Gene | No. of patho-<br>genic<br>variants | No. of<br>patients | No. of<br>families | Range of<br>age of onset<br>(years) | CoQ<sub>10</sub> level<br>(% of control) |  | Common clinical<br>manifestations <sup>a</sup> | No. of<br>CoQ<sub>10</sub>-<br>treated<br>patients <sup>b</sup> |
| --- | --- | --- | --- | --- | --- | --- | --- | --- |
|  |  |  |  |  | [No. of patients examined] |  |  |  |
|  |  |  |  |  | Skin fibroblasts | Muscle |  |  |
| *PDSS1* | 1 | 2 | 1 | 1-3 | <5% [2] | ND | encephalopathy,<br>deafness | 0 |
| *PDSS2* | 5 | 4 | 4 | infancy-2 | 12% [1] | 14% [1] | NS,<br>encephalopathy | 2 |
| *COQ2* | 20 | 25 | 17 | infancy-68 | 9-36% [7] | 3-38% [5] | NS,<br>encephalopathy | 10 |
| *COQ4* | 27 | 32 | 25 | infancy-9 | 22-98% [8] | 2-63% [6] | encephalopathy,<br>cardiac pathology | 21 |
| *COQ5* | 1 | 3 | 1 | childhood | ND | 57% [1] | cerebellar ataxia,<br>encephalopathy | 3 |
| *COQ6* | 15 | 28 | 20 | infancy-6.4 | ND | ND | NS, SND | 5<sup>2</sup> |
| *COQ7* | 7 | 6 | 5 | infancy-<br>childhood | 10-70% [4] | 10% [1] | spasticity, motor<br>difficulties | 3 |
| *COQ8A/*<br>*ADCK3* | 79 | 112 | 88 | infancy-42 | 35-100% [8] | 2-100% [13] | cerebellar ataxia,<br>exercise<br>intolerance | 59<sup>2</sup> |
| *COQ8B/*<br>*ADCK4* | 36 | 88 | 51 | infancy-32 | 27% [2] | ND | NS | 33 |
| *COQ9* | 3 | 3 | 3 | infancy | 11-18% [2] | 15% [1] | encephalopathy | 2 |

ND: not determined; NS: nephrotic syndrome; SND: sensorineural deafness. <sup>a</sup> The most common symptoms among the reported patients are listed. <sup>b</sup> Including treatment with ubiquinol (reduced form of CoQ); in addition, the number of patients treated with the CoQ analogue idebenone are indicated as superscripts.

In the final analysis, we included and assessed a total of 89 patients. The results are shown in Table 2. Details, including total count of patients treated and numbers of exclusions for each gene, can be found in Table S3. We classified 65 out of the 89 patients (73.0%) as not responding to CoQ_10_ treatment according to the evaluation criteria (Table S4). Among those, there are nine cases in which patients showed infantile onset and multisystem involvement. Such cases may be more challenging to treat, but this is only speculation. Of the 24 cases (27.0%) that were identified as responders, 20 were found to provide objective descriptions of responses and four are considered to be responders because they meet the criterion of having a subjective description of responses to CoQ_10_ therapy (Table S5). Note, however, that all responses were partial, and responses are frequently only observed with a single symptom. Table 3 highlights the five cases in which a worsening of patient conditions after stopping/halting of CoQ_10_ treatment or regimen change was reported. Four out of the five also reported recovery to some extent following treatment resumption. These cases potentially provide the most tantalizing evidence for a partial efficacy of CoQ_10_ treatment for CoQ deficiency. We should note, however, that the possibility of placebo effects cannot be excluded. Furthermore, in one of the cases the patient’s condition was reported to worsen after replacement of CoQ_10_ with idebenone and thus it is impossible to distinguish between the effects of stopping CoQ_10_ and potential idebenone toxicity. Of the other 15 cases of responses with objective description, four cases reported a decrease of proteinuria after CoQ_10_ treatment as an indication of kidney function improvement and ten reported a reduction in a severity score of ataxia or another motor performance test at a follow-up. However, five of the patients classified as responders because of an amelioration of proteinuria had only kidney symptoms, and in two cases only proteinuria.

**Table 2.** Reported partial effects of CoQ_10_ treatment in primary CoQ deficiency patients.

| Gene | No. of patients included in the analysis | Responding |  | Not responding |
| --- | --- | --- | --- | --- |
|  |  | Objective description | Subjective description |  |
| *PDSS1* | 0 | - | - | - |
| *PDSS2* | 2 | 0 | 0 | 2 |
| *COQ2* | 7 | 1 | 0 | 6 |
| *COQ4* | 19 | 3 | 2 | 14 |
| *COQ5* | 3 | 3 | 0 | 0 |
| *COQ6* | 3 | 1 | 0 | 2 |
| *COQ7* | 3 | 0 | 0 | 3 |
| *COQ8A/ADCK3* | 41 | 9 | 2 | 30 |
| *COQ8B/ADCK4* | 9 | 3 | 0 | 6 |
| *COQ9* | 2 | 0 | 0 | 2 |

**Table 3.**
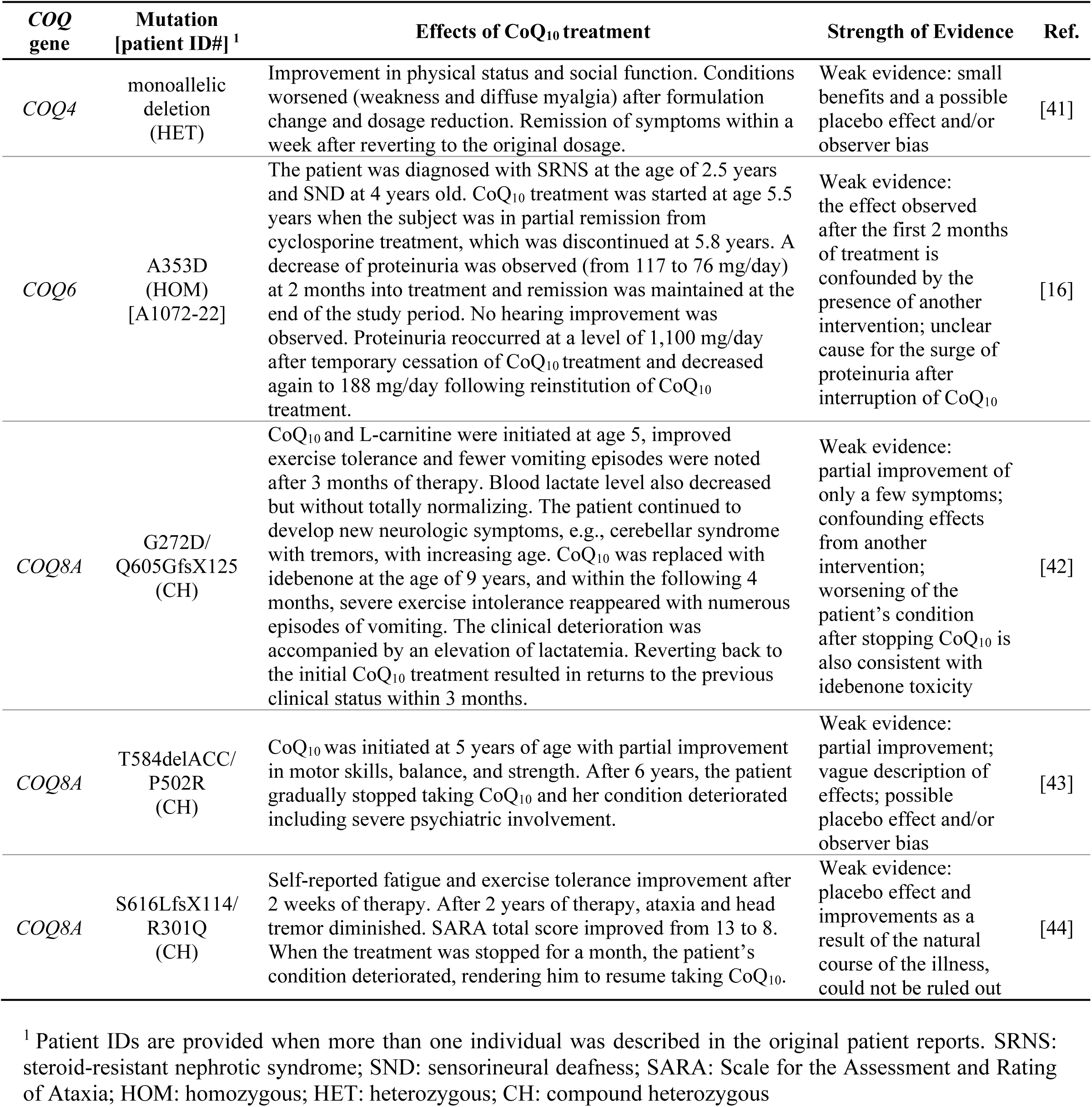
Therapeutic efficacy of CoQ_10_ suggested by the effects of treatment interruptions.

Treatment effects established by quantitative or semi-quantitative measures to describe the response to CoQ_10_ treatment were counted as responding with objective description, while descriptions of positive effects but without relying on a quantitative or semi-quantitative measures were counted as responding with subjective description. “Not responding” include the patients who were reported not to respond to CoQ_10_ treatment or whose responses we consider lacking a convincing demonstration of a response to CoQ_10_ supplementation.

As shown in Fig. 3 and S1, there is no significant differences in treatment dosage and duration of treatment between the non-responding and responding patients. The highest reported dosage is 2,100mg/day. No substantial adverse effects have been reported for the CoQ_10_-treated PCoQD patients. However, an adverse reaction has been reported in one case of treatment with the synthetic CoQ analogue idebenone which has a hydroxydecyl instead of a decaprenyl side chain and higher solubility than CoQ_10_ [45].

**Figure 3.**
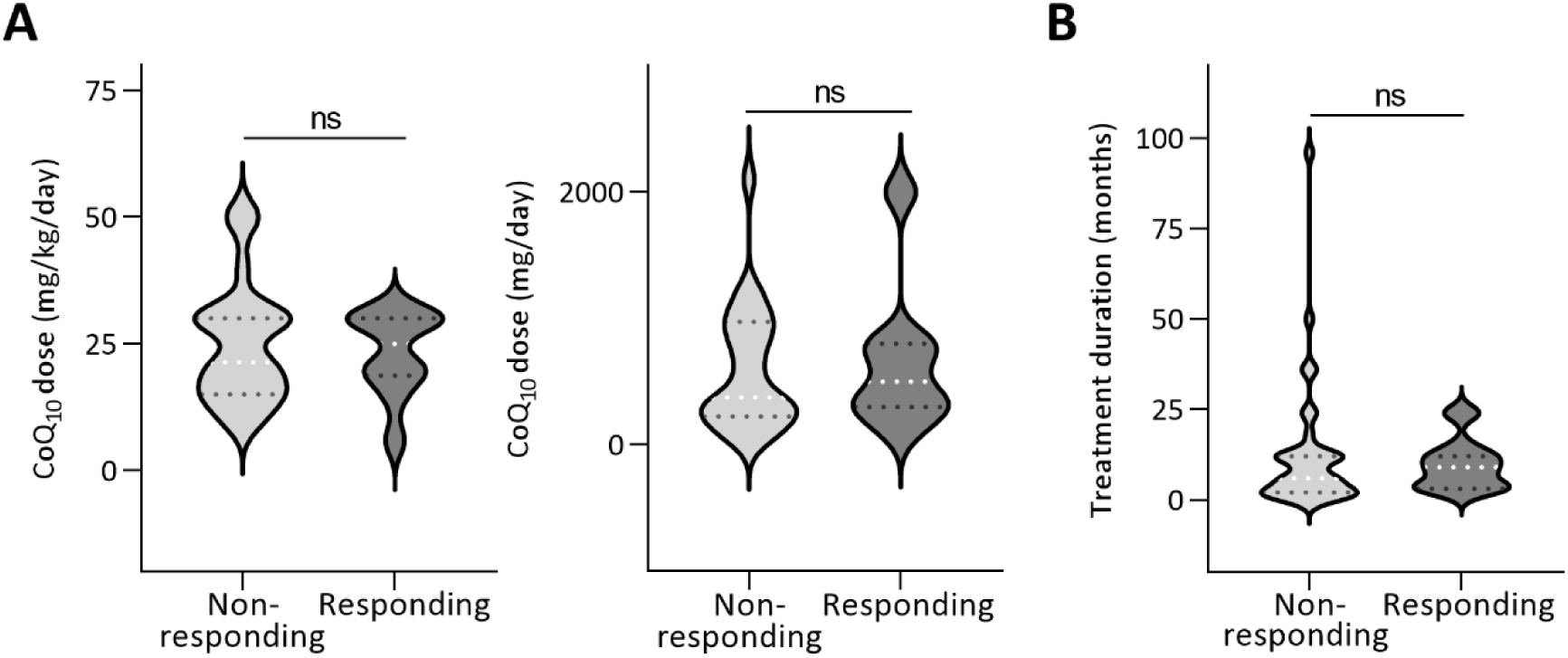
The violin plots of CoQ_10_ treatment dose and duration. Two graphs are shown for dosage comparison because CoQ_10_ treatment dosages were reported in 2 different units. ns: not significant (Student’s *t*-test).

## DISCUSSION

In humans, mutations have so far been reported in all the genes required for CoQ_10_ biosynthesis, except *COQ3*. COQ3 is an O-methyltransferase and it is the only COQ protein that is required for more than one step in the CoQ biosynthetic pathway [46]. Thus, one possible explanation for the lack of reports of COQ3 patients is that, because it is required for two enzymatic steps, pathogenic mutations in *COQ3* are more detrimental to CoQ production and thus are more likely to be lethal. Among the reported PCoQD patients, 37.0% (112/303) carry a mutation in the *COQ8A* gene and 29.0% (88/303) carry a mutation in the *COQ8B* gene. The reason for the higher *COQ8A* and *COQ8B* patient counts is most likely because genetic screening studies were performed for *COQ8A* and *COQ8B* on a relative larger scale. Two studies reported screening for *COQ8A* mutations in patients with ataxic symptoms, resulting in the identification of 69 patients carrying rare biallelic variants [15, 39]. Screens for *COQ8B* mutations in patients with renal disorders, including nephrotic syndrome and chronic renal failure, were described in three studies, which in total reported the identification of 63 *COQ8B* patients [47-49]. COQ8A and COQ8B are orthologs of yeast Coq8p, which plays a regulatory role in CoQ biosynthesis [40, 47, 50]. COQ8A is expressed in most tissues, but there is a relative enrichment of COQ8B in podocytes [47, 51]. Consistently, *COQ8B* patients were described to have a less severe clinical course and manifest largely kidney-limited phenotypes [47, 52]. In mice, the *Coq8a*^−*/*−^ model was shown to develop ataxia accompanied by minor neurological and muscle phenotypes [51]. More interestingly, unlike for other *Coq* genes, including *Coq8b*^*-/-*^, which are embryonically lethal, *Coq8a*^−*/*−^ mice are viable and maintain a moderate level of residual CoQ [51, 53-59]. Thus, the mutation frequency observed for a given *COQ* gene is likely influenced by the role it plays in CoQ biosynthesis and its tissue expression pattern. With increasing affordability and accessibility of genome or exome sequencing [60], more and more PCoQD patients are being reported, and a more accurate picture of PCoQD patients’ frequency should soon emerge.

Often CoQ_10_ deficiency patients are started on oral CoQ_10_ supplementation immediately after diagnosis. Various oral formulations of CoQ_10_ are available [61]. The scientific literature as well as the general media mostly state that oral CoQ_10_ supplementation is effective and thus that CoQ_10_ deficiency is treatable [33]. However, to the best of our knowledge, there is no other evidence that could support such a belief than the set of studies reviewed here. The final step of our analysis is based on published studies on 89 PCoQD patients for which we consider there to be sufficient information available to estimate the clinical effectiveness of the CoQ_10_ treatment. Of them, 65 cases fit our criteria for not-responding, including patients with age of onset ranging from neonatal to 42 years of age and that present with multi-system symptoms or primarily one organ-specific manifestation (e.g., cerebellar ataxia or nephrotic syndrome). Among the 24 cases identified as responsive, 12 cases reported improvement of an ataxia rating score and 7 out of them are patients with *COQ8A* mutations for whom ataxia is often the most prominent symptom. Five cases reported proteinuria improvement at a post-treatment follow-up, and in all five of them renal dysfunction was the only manifestation. However, many PCoQD patients with ataxia or kidney symptoms were reported to show no response or the condition continued to deteriorate after CoQ_10_ treatment (Table S4). Therefore, the observed relative prevalence of positive effects on ataxia or proteinuria does not indicate that the kidneys and cerebellum are more sensitive to supplemental treatment with CoQ_10_. Of note, none of the studied that reported symptomatic improvement found a profound rescue of the patients’ conditions. Furthermore, in patients with multisystem manifestations, effects were reported only for a few symptoms and most of the other symptoms still persisted after CoQ_10_ treatment. Detrimental effects of treatment interruption were noted in five cases, which potentially constitute the best evidence for some effectiveness of CoQ_10_ therapy. However, as these are not blinded studies, the possibility of placebo effects remains of concern.

Overall, most descriptions of the effects of CoQ_10_ treatment have incomplete information and lack a complete clinical picture. Doctors and patients are aware of the treatments (i.e., no blinding). There can of course be no “no-treatment” control group of patients. For these reasons, we consider the cases where a minimal effect only was reported as not responding to treatment. It has been hypothesized that CoQ treatment cannot reverse severe tissue damage due to PCoQD when the disease has already progressed too far before therapy is initiated [30, 62]. However, animal studies with an unnatural CoQ biosynthetic precursor suggest that most phenotypes due to severe CoQ deficiency can be completely rescued by a partial replenishment of CoQ levels [63-65]. Therefore, it is reasonable to expect that significant clinical benefits should be possible even in severely impaired PCoQD patients if in fact a significant amount of CoQ_10_ were absorbed and could reach affected tissues.

The results from our analysis indicate that most PCoQD patients treated with CoQ_10_ showed little or no response, and, in the cases of positive reports, the overall clinical benefit was only very limited. This strongly suggests a lack of efficacy of CoQ_10_ treatment. It is noteworthy that clinical trials have been conducted to assess the potential benefit of CoQ_10_ in the treatment of patients with secondary CoQ_10_ deficiency or mitochondrial disease. CoQ_10_ supplementation was shown to elicit no benefit to the patients with statin-induced myalgia [66]. To date, only few double-blind and randomized clinical trials evaluating CoQ_10_ in the treatment of mitochondrial disorders have been completed. There were reports of minor effects for improved muscle strength and attenuation of lactate rise post-exercise. However, the overall conclusion remained that CoQ_10_ is ineffective for the treatment of patients with mitochondrial disorder, or at least there is no solid evidence to suggest otherwise [67, 68]. It is particularly worthy of mention that a recent phase three, randomized, double-blind, clinical trial evaluating CoQ_10_ in the treatment of children with primary mitochondrial respiratory disease reported no significant improvements in any of the measured clinical outcome variables (https://clinicaltrials.gov/NCT00432744).

CoQ_10_ is extremely lipophilic and practically insoluble in water; therefore, to develop pharmaceutical CoQ preparations, a number of formulation strategies for insoluble compounds have been tried, such as oil solution, emulsion, cyclodextrin complexation, and liposomal nanoencapsulation [69] Presently, all currently marketed formulations of CoQ_10_ are for oral administration only. Like all dietary lipids, orally administered CoQ_10_ is absorbed in the enterocytes, packaged into chylomicrons (large lipoprotein particles) and then transported via the lymphatics to the circulation (Fig. 1C) where CoQ_10_ is mostly packaged into lipoproteins [70]. In humans, the level of total plasma CoQ_10_ is less than 2 μg/mL. Increases several-fold above normal plasma level has been reported after CoQ_10_ treatment [70-72]. However, it is not known how blood CoQ_10_ concentration is related to effectiveness in relieving symptoms. Moreover, the mechanism of tissue uptake of CoQ_10_ is still poorly understood. In rodents, after oral CoQ_10_ supplementation high concentrations of CoQ_10_ were reported for several tissues including the liver, ovaries, brown adipocytes, and spleen after feeding CoQ_10_-supplemented food or water, but not for the heart, kidney, muscle and brain, the main affected tissues in PCoQD [63, 73-77]. Key factors that influence the tissue or cellular uptake of CoQ_10_ await future studies.

There have been discussions on the possible merits of using the reduced form of CoQ_10_, also known as ubiquinol, to enhance the bioavailability of CoQ_10_ [78]. However, this is not yet strongly supported by all studies, and ubiquinol’s claimed to superior bioavailability is still in question [70]. Out of the 89 cases included in our final analysis, 6 were reported to be treated with ubiquinol (Table S4 and S5). Two met our criteria of responding and 4 did not. Thus, this data also does not point to better bioavailability of ubiquinol over regular CoQ_10_ in PCoQD patients.

In sum, the results of the present review suggest the need to develop alternative strategies of providing CoQ_10_ and stresses the need for caution when seeking to justify the widespread use of CoQ_10_ for disease treatment or as a dietary supplement. Our recent study suggests the possibility of intravenously administering CoQ_10_ solubilized with the fungicide caspofungin to achieve much higher plasma concentration and thus more effective CoQ_10_ therapy [79]. Furthermore, modified precursors of the quinone ring of CoQ_10_, for example, DHB, have been considered as potential alternative treatment option for some types of PCoQD [63-65, 80, 81]. Future work is warranted to further explore these possibilities and unleash the full potential of CoQ_10_ therapy.

## Data Availability

All data produced in the present work are contained in the manuscript.

## AUTHOR CONTRIBUTION

SH and YW designed the study. YW did literature searches and extracted data. SH verified data accuracy, narrative summaries, and interpretations. Both authors contributed to the selection of included studies, evaluation of data quality, and data analyses. SH and YW wrote the manuscript together and approved the final manuscript.

## SUPPLEMENTARY MATERIAL

Table S1: Primary CoQ_10_ deficiency patients identified by literature search.

Table S2: Cases excluded from the final analysis and reasons for their exclusion.

Table S3: Partial effects reported for CoQ_10_ treatment of primary CoQ deficiency patients.

Table S4: Patient cases classified as not responding to CoQ_10_ treatment.

Table S5: Cases with positive outcomes following CoQ_10_ treatment, classified as responding.

Fig. S1: The violin plot of total CoQ_10_ amounts taken.

