## SUPPLEMENTARY MATERIAL for "The efficacy of coenzyme Q_10_ treatment in alleviating the symptoms of primary coenzyme Q_10_ deficiency: a systematic review"

### Supporting Information

#### The search query used in PUBMED

((((((((((((((((COQ2[Title/Abstract]) OR (PDSS1[Title/Abstract])) OR (PDSS2[Title/Abstract])) OR (COQ3[Title/Abstract])) OR (COQ4[Title/Abstract])) OR (COQ[Title/Abstract])) OR (COQ6[Title/Abstract])) OR (COQ7[Title/Abstract])) OR (COQ8A[Title/Abstract])) OR (ADCK3[Title/Abstract])) OR (COQ8B[Title/Abstract])) OR (ADCK4[Title/Abstract])) OR (COQ9[Title/Abstract])) OR (COQ10[Title/Abstract])) AND (patient[Title/Abstract]) ) AND (primary[Title/Abstract])

**Table S1 Primary CoQ10 deficiency patients identified by literature search.**

**S1.1 Primary CoQ$_{10}$ deficiency-2 (COQ10D2; 614651) due to mutations in the *PDSS1* gene [# of patients: 2]**

| Gene [Patient ID] | # of patients (# of families) | Mutation | Level of CoQ$_{10}$ (% of control) | Age at onset (sex) if known | Symptoms | Biochemical tests and muscle pathology | RCC enzymes | CoQ$_{10}$ dose and responses | Age at last reported exam or death | Corresponding PI | Reference |
|---|---|---|---|---|---|---|---|---|---|---|---|
| **PDSS1 [1]** | 2 (1) | D308E (HOM) | <5% (fibroblasts) | 1-3 y/o (M) | encephalopathy, deafness, cardiac valvulopathy, livedo reticularis, mild mental retardation, macrocephaly, peripheral neuropathy, bulimia, obesity, and optic atrophy | mildly elevated blood lactate, mitochondrial aggregates in the muscle | CII+CIII ↓, G3PDH+CIII↓, CII+CIII/CS↓ (fibroblasts), CI+CIII/CI↓, CIII/CI+CIII ↑ (muscle mitochondria) | no data | 22 years of age | Agnès Rötig, Hôpital Necker-Enfants Malades, France | (Mollet et al., 2007) |
| **PDSS1 [2]** | | D308E (HOM) | <5% (fibroblasts) | 2 y/o (F) | deafness, cardiac valvulopathy, obesity, macrocephaly, optic atrophy, peripheral neuropathy, livedo reticularis, mental retardation | mildly elevated blood lactate | no data | no data | 14 years of age | | |

**S1.2 Primary CoQ$_{10}$ deficiency-3 (COQ10D3; 614652) due to mutations in the *PDSS2* gene [# of patients: 4]**

| Gene [Patient ID] | # of patients (# of families) | Mutation | Level of CoQ$_{10}$ (% of control) | Age at onset (sex) if known | Symptoms | Biochemical tests and muscle pathology | RCC enzymes | CoQ$_{10}$ dose and responses | Age at last reported exam or death | Corresponding PI | Reference |
|---|---|---|---|---|---|---|---|---|---|---|---|
| **PDSS2 [1]** | 1 (1) | Q332X/ S382L (CH) | ~ 14% (muscle) ~ 12% (fibroblasts) | 3 m/o (M) | NS, hypotonia, Leigh syndrome, seizure | blood lactate↑, serum albumin↓, proteinuria mitochondrial aggregates in muscle | CII+CIII↓ (fibroblasts and muscle) | 50mg/day beginning at age 3 months, no response *[NR]* | died at age of 8 months | Michio Hirano, Columbia University medical center, USA | (Lopez et al., 2006; Quinzii et al., 2008; Salviati et al., 2012) |
| **PDSS2 [2]** | 1 (1) | S382L (HOM) | no data | 1.9 y/o (M) | SRNS, cerebral palsy, intellectual disability | no data | no data | no data | no data | Friedhelm Hildebrandt, | |

| | | | | | | | | | | | |
|---|---|---|---|---|---|---|---|---|---|---|---|
| **PDSS2 [3]** | 1 (1) | A384D (HOM) | no data | neonatal (M) | SRNS | no data | no data | no data | no data | Boston Children's Hospital, USA | (Sadowski et al., 2015) |
| **PDSS2 [4]** | 1 (1) | H162R/ c.1042_1148-2816del (CH) | no data | neonatal (M) | NS, encephalomyopathy, hypertrophic cardiomyopathy, deafness, retinitis pigmentosa, global developmental delay | blood lactate↑, proteinuria | no data | 20mg/kg/day, no response *[NR]* | died at age of 8 months (1 month after admission) | Béla Iványi, University of Szeged, Hungary | (Ivanyi et al., 2018) |

### S1.3 Primary $CoQ_{10}$ deficiency-1 (COQ10D1; 607426) due to mutations in the *COQ2* gene [# of patients: 25]

| Gene [Patient ID] | # of patients (# of families) | Mutation | Level of $CoQ_{10}$ (% of control)[1] | Age at onset (sex) if known | Symptoms | Biochemical tests and muscle pathology | RCC enzymes | $CoQ_{10}$ dose and responses | Age at last reported exam or death | Corresponding PI | Reference |
|---|---|---|---|---|---|---|---|---|---|---|---|
| **COQ2 [1]** | 1 (1) | R197H/ N228S (CH) | ~ 36% (fibroblasts) <3% (kidney, muscle) | 18 m/o (M) | SRNS | normal blood lactate | CII+CIII ↓ (muscle) | 30mg/kg/day since age 21 months, response not described | 29 months of age, ESRF at age 20 months | Francesca Diomedi-Camassei, Bambino Gesu` Children's Hospital, Italy | (Diomedi-Camassei et al., 2007; Quinzii et al., 2010) |
| **COQ2 [2]** | 1 (1) | S146N (HOM) | ~ 17% (fibroblasts) <3% (kidney, muscle) | neonatal (M) | glomerulonephritis, acute renal failure, seizure, epileptic encephalopathy | CSF lactate↑ | CII+CIII↓ (muscle) | no data | died at 6 months of age | Francesca Diomedi-Camassei Bambino Gesu` Children's Hospital, Italy | (Diomedi-Camassei et al., 2007) (Bujan et al., 2014) |
| **COQ2 [3]** | 2 (1) | Y297C (HOM) | ~18% (fibroblasts) ~ 37.5% (muscle) | 11 m/o (M) | infantile encephalomyopathy, SRNS/FSGS, hypotonia, optic atrophy, tremors, psychomotor regression | normal lactate levels, proteinuria; myofibers with excessive succinate dehydrogenase staining | CI+CIII ↓, CII+CIII ↓ (muscle), CII+CIII ↓ (fibroblasts) | 30 mg/kg/day beginning at age 22months, neurologic picture improved, but no change in renal function *[NR]* | ESRF at age 18 months, kidney transplant at age of 3 years | Michio Hirano, Columbia University medical center, USA | (Diomedi-Camassei et al., 2007; Montini et al., 2008; Quinzii et al., 2006; Quinzii et al., 2008; Salviati et al., 2005) |
| **COQ2 [4]** | | Y297C (HOM) | ~ 17% (fibroblasts) | 12 m/o (F) | NS/FSGS without any clinical signs of neurologic involvement. | proteinuria. hypoalbuminemia | CII+CIII ↓ (fibroblasts) | △30 mg/kg/day, there was no improvement during the first 2 weeks of treatment; an episode of acute renal failure required continuous hemofiltration for | 5 years of age | Michio Hirano, Columbia University medical center, USA | (Diomedi-Camassei et al., 2007; Montini |

| | | | | | | | | | | | |
|---|---|---|---|---|---|---|---|---|---|---|---|
| | | | | | | | | 4 days. 20 days after the initiation of the treatment, recovery of renal function and a reduced level of proteinuria was observed. After 50 months of therapy, renal function remains normal, though proteinuria was still present (other medication: diuretics) *[NR]* | | | et al., 2008; Quinzii et al., 2006; Quinzii et al., 2008; Salviati et al., 2005) |
| **COQ2 [5]** | 1 (1) | N401fsX 415 (HOM) | ~ 24% (fibroblasts) | neonatal (F) | Infantile multiorgan failure (neurologic distress, liver failure, NS, anemia, pancytopenia, insulin-dependent diabetes, and seizures) | no data | CI+CIII ↓, CII+CIII ↓ (liver) | no data | died at 12 days | Agnès Rötig, Hôpital Necker-Enfants Malades, France | (Mollet et al., 2007; Quinzii and Hirano, 2010) |
| **COQ2 [6]** | 1 (1) | L234fsX2 47/ N228S (CH) | no data | 2 y/o (F) | SRNS | no data | no data | no data | 4 years of age | Moin A. Saleem, Southmead Hospital, UK | (McCarth y et al., 2013) |
| **COQ2 [7]** | 2 (1) | A302V (HOM) | ~ 29.2% (muscle) | neonatal (F) | generalized edema, seizures, apnea, hypotonia, dystonic-hyper kinetic movement, feeding problems | blood lactate↑ | CII+CIII↓ (muscle, fibroblasts) | no data | died at age of 5 months | R.J.T. Rodenburg, Radboud University, The Netherlands | (Jakobs et al., 2013) |
| **COQ2 [8]** | | A302V (HOM) | ~ 8.5% (fibroblasts), 3.4% (muscle) | neonatal (M) | generalized edema, seizures, apnea, hypotonia, dystonic-hyper kinetic movement, feeding problems | blood lactate↑, muscle histology is normal | CI↓, CIII↓, CII+CIII↓, ATP↓ (muscle), CII+CIII↓(fibroblast s) | no data | died at age of 6 months | R.J.T. Rodenburg, Radboud University, The Netherlands | (Jakobs et al., 2013; Ziosi et al., 2017) |
| **COQ2 [9]** | 1 (1) | S109N (HOM) | ~ 11.4% (fibroblasts) | neonatal (M) | peripheral hypertonia, cardiomyopathy, hypertrophic cardiomegaly, nephrotic syndrome | CSF lactate ↑, proteinuria | CII+CIII↓(kidney) | 30mg/kg/day, no response *[NR]* | died at age of 5 months | Emmanuel Scalais, Centre Hospitalier de Luxembourg, Luxembourg | (Scalais et al., 2013; Ziosi et al., 2017) |
| **COQ2 [10]** | 1 (1) | S146N/ R387X (CH) | no data | neonatal (F) | acidosis, hyperglycemia, cardiomegaly, respiratory distress, necrotizing enterocolitis, encephalopathy | blood lactate↑ | CI↓, CII↓, CS↑ (muscle) | no data | died at age of 2 months | D. Dinwiddie, University Of New Mexico, USA | (Dinwidd ie et al., 2013) |
| **COQ2 [11]** | 2 (1) | M128V-V393A (HOM) | <20% (brain) | 68 y/o (F) | multiple system atrophy with predominant parkinsonism, retinitis pigmentosa | no data | no data | no data | died | Shoji Tsuji, University of Tokyo Japan | (Multiple -System Atrophy Research, 2013) |
| **COQ2 [12]** | | M128V-V393A (HOM) | no data | 62 y/o (M) | multiple system atrophy with predominant parkinsonism, ataxia, retinitis pigmentosa | no data | no data | no data | died | | |
| **COQ2 [13]** | 2 (1) | M387X/ V393A (CH) | no data | 50 y/o (F) | multiple system atrophy of the cerebellar type | no data | no data | no data | no data | | |

| | | | | | | | | | | |
|---|---|---|---|---|---|---|---|---|---|---|
| **COQ2 [14]** | | M387X/ V393A (CH) | no data | 44 y/o (M) | multiple system atrophy of the cerebellar type | no data | no data | no data | no data | | |
| **COQ2 [15]** | 1 (1) | N228S (HOM) | no data | no data (M) | SRNS | no data | no data | no data | no data | Friedhelm Hildebrandt, Boston Children's Hospital, USA | Sadowski et al., 2015) |
| **COQ2 [16]** | 1 (1) | R173H/ N228S (CH) | no data | 2.5 y/o (M) | SRNS | no data | no data | no data | no data | | |
| **COQ2 [17]** | 1 (1) | N228S/ L286F (CH) | no data | 1.3 y/o (F) | SRNS | no data | no data | no data | no data | | |
| **COQ2 [18]** | 1 (1) | Y297C (HOM) | no data | 5 m/o (M) | SRNS | no data | no data | no data | no data | | |
| **COQ2 [19]** | 2 (1) | G390A (HOM) | no data | 18 y/o (F) | SRNS/FSGS | dysmorphic mitochondria (kidney) | no data | treated, response not described | kidney transplant at age 20 years | L Gesualdo, University "Aldo Moro", Italy | (Gigante et al., 2017) |
| **COQ2 [20]** | | G390A (HOM) | no data | 16 y/o (F) | SRNS/FSGS | dysmorphic mitochondria (kidney) | no data | treated, response not described | kidney transplant at age 19 years | | |
| **COQ2 [21]** | 3 (1) | c.288dup C/ R126G (CH) | no data | 25 y/o (M) | diffuse glomerulosclerosis, end-stage nephropathy, retinopathy | no data | no data | 30 mg/kg/day for 6 months, no ERG improvement, but best corrected visual acuity and areas of retinal atrophy on autofluorescence were noted to be stable on treatment *[NR]* | 25 years of age | Stephen H. Tsang, Columbia University Irving Medical Center, USA | (Abdelhakim et al., 2020) |
| **COQ2 [22]** | | c.288dup C/ R126G (CH) | no data | 21 y/o (M) | mesangial sclerosis, end-stage nephropathy, retinopathy, lymphoma | no data | no data | | 32 years of age, kidney transplant at age 5 years | | |
| **COQ2 [23]** | | c.288dup C/ R126G (CH) | no data | 23 y/o (F) | retinopathy, end-stage nephropathy | no data | no data | | 28 years of age, kidney transplant at age 10 years | | |
| **COQ2 [24]** | 2 (1) | Y353C/ T325A (CH) | no data | 2 y/o (M) | SRNS/FSGS | no data | no data | no data | died of ESRF at 5 years of age | Liangzhong Sun, Southern Medical University, China | (Li et al., 2021) |
| **COQ2 [25]** | | Y353C/ T325A (CH) | no data | 7 m/o (F) | SRNS | no data | no data | 30 mg/kg/ day beginning at age 11 months, urinary protein decreased with the increasing dose of $CoQ_{10}$, now on the dosage of 600mg/day *[Obj.]* | normal growth at 4 years old | | |

**S1.4 Primary $CoQ_{10}$ deficiency-7 (COQ10D7; 616276) due to mutations in the *COQ4* gene [# of patients: 32]**

| Gene [Patient ID] | # of patients (# of families) | Mutation | Level of $CoQ_{10}$ (% of control)[1] | Age at onset (sex) if known | Symptoms | Biochemical tests and muscle pathology | RCC enzymes | $CoQ_{10}$ dose and response | Age at last reported exam or death | Corresponding PI | References |
|---|---|---|---|---|---|---|---|---|---|---|---|

| | | | | | | | | | | | |
|---|---|---|---|---|---|---|---|---|---|---|---|
| **COQ4 [1]** | 1 (1) | mono-allelic deletion (CH) | ~ 43% (fibroblasts) | neonatal (M) | dysmorphic features, mental retardation, encephalomyopathy | blood lactate in normal range, increased SDH staining in the muscle | CII+CIII↓ (fibroblasts) | 30 mg/kg/day, improvement in physical status and social function. Conditions worsened (weakness and diffuse myalgia) after formulation change and dosage reduction to 2mg/kg/day. Remission of symptoms within a week after reverting back to the original dosage. Then switched to 15mg/kg/day of ubiquinol *[Obj.]* | 3 years of age | Plácido Navas, Universidad Pablo de Olavide, Spain | (Salviati et al., 2012) |
| **COQ4 [2]** | 1 (1) | R145G (HOM) | ~ 41-54% (fibroblasts), ~ 23% (muscle) | neonatal (M) | hypotonia, areflexia, acrocyanosis, bradycardia, respiratory insufficiency, left ventricular hypoplasia | blood lactate↑, blood creatine kinase↑ | CI+CIII↓, CII+CIII↓, CI↓ (autoptic muscle), CII+CIII↓ (fibroblasts) | not treated | died at 4 hours after birth | Holger Prokisch, Technische Universität München, Germany | (Brea-Calvo et al., 2015; Ziosi et al., 2017) |
| **COQ4 [3]** | 1 (1) | R141X/ G240C (CH) | no data | neonatal (F) | respiratory failure, lactic acidosis, cardiomyopathy, heart failure | urinary and plasmatic amino acids, organic acids, and acylcarnitine are normal | CI ↓, CII ↓, CIII ↓, CIV ↓, CI+CIII ↓ (autoptic muscle) | not treated | died at 4 hours after birth | Holger Prokisch, Technische Universität München, Germany | (Brea-Calvo et al., 2015) |
| **COQ4 [4]** | 2 (1) | L52S/ T174del (CH) | ~ 2% (muscle) | neonatal (F) | distal arthrogryposis, respiratory distress, encephalopathy, multiorgan failure | blood lactate↑ | CIV↓, CII+CIII↓ (autoptic muscle) | not treated | died at 3 days after birth | | |
| **COQ4 [5]** | | L52S/ T174del (CH) | ~ 3% (muscle) | neonatal (F) | respiratory distress, encephalopathy | blood lactate↑, amino acids in plasma↑, analysis of urinary organic acids showed mitochondrial dysfunctional excretion pattern | CIII↑, CIV↑ (autoptic muscle) | not treated | died at 2 days after birth | | |
| **COQ4 [6]** | 1 (1) | P64S (HOM) | ~ 63% (muscle) | 10 m/o (M) | motor deterioration, ataxia, epileptic seizures, swallowing impairment, progressive scoliosis, cognitive deterioration | blood tests excluded liver and kidney involvement and showed no lactic acidosis | CI↓, CIII↓, CI+CIII ↓ (muscle) | treated, response not described | 17 years of age | | |
| **COQ4 [7]** | 2 (1) | L82Q/ R158Q (CH) | ~ 16% (muscle) | neonatal (F) | seizures, severe lactic and respiratory acidosis, heart failure | blood and CSF lactate↑, plasma alanine↑, increased mitochondrial size in the muscle | CII+CIII↓ (muscle) | △20 mg/kg/day beginning at the first day of life, which resulted in normalization of lactate and improvement in cardiac function. Nevertheless, the patient continued exhibiting | died at 2 months of age | Marwan Shinawi, Washington University School of Medicine, USA | (Chung et al., 2015) |

| | | | | | | | | | | | |
|---|---|---|---|---|---|---|---|---|---|---|---|
| | | | | | | | | intermittent episodes of lactic acidemia and cardiac decompensation until death (other medications: thiamine, riboflavin, hydroxocobalamin, biotin) *[NR]* | | | |
| **COQ4 [8]** | | not tested | no data | neonatal (F) | respiratory distress, metabolic acidosis, apnoeic/gasping episode | no data | no data | not treated | died at 36 hours of life | | |
| **COQ4 [9]** | 1 (1) | R240C (HOM) | no data | neonatal (F) | hypotonia, cardiomyopathy, cerebellar and brainstem hypoplasia | lactic and pyruvic aciduria | normal ETC complex activities (muscle) | not treated | died at 4 days of life | | |
| **COQ4 [10]** | 2 (1) | R66Q/ D68H (CH) | no data | neonatal (F) | seizure, respiratory distress, intractable epilepsy, hypotonia, feeding difficulties, cardiomyopathy, and global developmental delay | blood lactate in normal range, a slight increase of CSF lactate | no data | no data | died at age of 19 months | | |
| **COQ4 [11]** | | not tested | no data | neonatal (F) | hypotonia, metabolic acidosis, | blood lactate↑, urinary malate and fumarate↑ | no data | no data | died at age of 10 weeks | | |
| **COQ4 [12]** | 1 (1) | R240C (HOM) | no data | neonatal (F) | poor/absent reflexes, cardiac hypertrophy, left hip dysplasia, hypotonia, episodes of apnea and bradycardia | normal lactate, pyruvate, ammonia, creatine phosphokinase, acylcarnitine and plasma amino acids, increased lactic acid, 2-ketoglutaric acid, fumarate and 2-hydroxyglutaric acid in urine, CSF lactate↑ | no data | △15 mg/kg/day beginning at age 1 month, no response [other medications: pyridoxal phosphate, folinic acid, and riboflavin] *[NR]* | died at 7 weeks old | | |
| **COQ4 [13]** | 1 (1) | V8AfsX19/D111Y + P119L (CH) | ~ 21% (muscle) ~ 34% (fibroblasts) | neonatal (M) | seizure, ventricular hypertrophy, bilateral hearing loss, hypotonia | blood lactate↑ | CI+CIII↓, CII+CIII↓ (fibroblasts) | no data | died at 4 months old | Ali B. Naini, Columbia University Medical Center, USA | (Sondheimer et al., 2017) |
| **COQ4 [14]** | 2 (1) | T77I (HOM) | no data | 4 y/o (M) | tremors, dysarthria, seizure, spastic tetraparesis and ataxia | no data | no data | 1000mg/day beginning at age 13, the 6 min walk test was stable over the period of a year *[NR]* | 15 years of age | Jan-Maarten Cobben, University of Amsterdam, the Netherlands | (Bosch et al., 2018) |
| **COQ4 [15]** | | T77I (HOM) | ~ 22% (fibroblasts) | 9 y/o (F) | seizure, dysarthria, spastic tetraparesis, ataxia | general laboratory tests were normal | no data | 1000mg/day beginning at age 11, the 6 min walk test was stable over a year, developed a second stroke-like episode at age 14 *[NR]* | 14 years of age | | |

| | | | | | | | | | | | |
|---|---|---|---|---|---|---|---|---|---|---|---|
| **COQ4 [16]** | 2 (1) | G124S (HOM) | no data | neonatal (M) | motor deterioration, weak responsiveness, hearing impairment, dystonia, seizure, tachycardia, respiratory distress | blood lactate↑, glucose↑, blood ammonia↑, no evidence of renal impairment | no data | no data | died at 5.6 months | Qiwei Guo, Xiamen University, China | (Lu et al., 2019) |
| **COQ4 [17]** | | G124S (HOM) | ~ 50% (fibroblasts) | neonatal (F) | motor deterioration, weak responsiveness, dystonia, nystagmus, respiratory distress, seizure | blood lactate↑, glucose↑, blood ammonia↑ | CII+CIII↓ (fibroblasts) | △50 mg/kg/day, improvement in seizure, screaming, and respiratory distress, no improvement in nystagmus, dystonia, psychomotor development, and ambulation *[NR]* | 1 year of age | | |
| **COQ4 [18]** | 1 (1) | P193S/ R240C (CH) | ~ 95% (fibroblasts) | 2.5 y/o (M) | developmental delay, hypotonia, sialorrhea, spasticity, ataxia | no data | no change of CII+CIII activity (fibroblasts) | 30 mg/kg/day of ubiquinol, improvement in neuromuscular symptoms after 2 months, further improvement of motor skills in the following months, but speech delay and cognitive impairment persisted *[Subj.]* | 2.7 years of age | Maria Marchese, IRCCS Fondazione Stella Maris, Italy | (Mero et al., 2021) |
| **COQ4 [19]** | 1 (1) | G95D/ R102H (CH) | ~ 98% (fibroblasts) | 5 y/o (F) | cognitive impairment, dysmetria, spastic ataxia, seizure | no data | no change of CII+CIII activity (fibroblasts), normal RCC activities (muscle) | 100mg/kg/day of ubiquinol, no response after 6 months (as assessed by the SARA scale) *[NR]* | 19 years of age | | |
| **COQ4 [20]** | 2 (1) | G55V (HOM) | normal range (blood) | 8 y/o (M) | ataxia, spasticity, epilepsy, cognitive deterioration, dysarthria, dysmetria and dysdiadochokinesia | no data | no data | 2000 mg/day, improvement of SARA score, dysarthria is persistent *[obj.]* | 27 years of age | Margit Burmeister, University of Michigan, USA | (Caglayan et al., 2019) |
| **COQ4 [21]** | | | normal range (blood) | 8 y/o (F) | dysarthria, spastic ataxia, epilepsy, cognitive deterioration, dysmetria, dysdiadochokinesia | no data | no data | Treated, dose not described, improvement of SARA score, gait difficulty and dysarthria are persistent *[obj.]* | 28 years of age | | |
| **COQ4 [22]** | 1 (1) | E161D (HET) | ~ 25% (fibroblasts) | 4 y/o (F) | mental retardation, rhabdomyolysis | muscle damage, rhabdomyolysis, disorganized intermyofibrillar pattern, SDH and COX staining↓ in the muscle | CI+CIII↓, CII+CIII ↓ (fibroblasts) | no data | 4 years of age | Pablo Menendez, CIBERONC, Spain | (Romero-Moya et al., 2017) |
| **COQ4 [23]** | 1 (1) | G124S/ c.402+1 G>C (CH) | low (fibroblasts) | neonatal (M) | encephalopathy, cardiomyopathy, visual and hearing impairment, respiratory failure, apnea, developmental delay | blood lactate↑ | CII+CIII↓ (fibroblasts) | 40 mg/kg/day beginning at 5 months of age, poor response *[NR]* | died at 8 months of age | Brian Hon-Yin Chung, Hong Kong Children's Hospital, China | (Yu et al., 2019) |
| **COQ4 [24]** | 1 (1) | G124S/ c.402+1 G>C (CH) | no data | neonatal (M) | cardiomyopathy, respiratory distress, metabolic acidosis | blood lactate and alanine↑ | no data | △15 mg/kg/day, no response [other medication: carnitine] *[NR]* | died at 2.5 days of age | | |

| | | | | | | | | | |
|---|---|---|---|---|---|---|---|---|---|
| **COQ4 [25]** | 1 (1) | G124S (HOM) | no data | neonatal (F) | cardiomyopathy, seizure, developmental delay | blood lactate↑, hyperammonemia | no data | △ treated, dose not described, cardiac function improved gradually and normalized after 10 days [other medication: intravenous immunoglobulin] *[NR]* | 9 months of age |
| **COQ4 [26]** | 2 (1) | G124S/ c.402+1 G>C (CH) | no data | neonatal (F) | seizure, apnea, encephalopathy, cardiomyopathy | blood lactate↑ | no data | started at the age of 4 years and 5 months, dose not described, no response observed after 1 month of treatment *[NR]* | 4.5 years of age |
| **COQ4 [27]** | | | no data | 2 m/o (F) | seizure, respiratory distress, cardiomegaly | blood lactate↑ | no data | started at 1 year of age, dose not described, no response, passed away 1 month later *[NR]* | died at 1.1 years of age |
| **COQ4 [28]** | 1 (1) | W184R/ c.402+1 G>C (CH) | low (fibroblasts) | 8 m/o (M) | microcephaly, developmental delay, dystonia, visual impairment, oro-motor dysfunction | blood lactate and alanine↑ | CII+CIII↓ (fibroblasts) | dose not described, no response *[NR]* | 3.6 years of age |
| **COQ4 [28]** | 1 (1) | G124S (HOM) | low (fibroblasts) | infancy (F) | visual impairment, dystonia, spasticity, developmental delay | blood lactate↑ | CII+CIII↓ (fibroblasts) | since age of 2 , dose not described, no response *[NR]* | died at 3.5 years of age |
| **COQ4 [30]** | 1 (1) | G124V/ G124S (CH) | low (fibroblasts) | infancy (F) | encephalopathy, dystonia, spasticity, developmental delay, visual impairment, seizure | blood lactate and alanine↑ | CII+CIII↓ (fibroblasts) | △beginning at 9 months of age, dose not described, subjective improvement in response [other medication: levetiracetam] *[NR]* | 3.3 years of age |
| **COQ4 [31]** | 1 (1) | G124S (HOM) | low (fibroblasts) | 2 m/o (M) | encephalopathy, spasms, seizure, development delay | blood lactate↑ | CII+CIII↓ (fibroblasts) | beginning at 7 years of age, dose not described, response not described | 8 years of age |
| **COQ4 [32]** | 2 (1) | G124S (HOM) | no data | 2 m/o (F) | hypotonia, developmental delay, bilateral cortical blinding, seizure, cardiomyopathy | blood lactate↑ | no data | 30mg/kg/day beginning at 11 months of age, some improvement in seizure control and development *[Subj.]* | 1.5 years of age |

#### S1.5 Primary $CoQ_{10}$ deficiency-9 (COQ10D9; 619028) due to mutations in the *COQ5* gene [# of patients: 3]

| Gene [Patient ID] | # of patients (# of families) | Mutation | Level of $CoQ_{10}$ (% of control)[1] | Age at onset (sex) if known | Symptoms | Biochemical tests and muscle pathology | RCC enzymes | $CoQ_{10}$ dose and response | Age at last reported exam or death | Corresponding PI | Reference |
|---|---|---|---|---|---|---|---|---|---|---|---|
| **COQ5 [1]** | 3 (1) | biallelic duplication of last 4 exons | ~ 57% (muscle) ~ 50% (leukocytes) | childhood (F) | ataxia, dysarthria, seizures, cognitive disability, behavioral problems, epilepsy, myoclonus, dysarthric cerebellar speech, dysmetria, mild tremors and mild lower limb spasticity | liver and renal function tests, carnitine and acyl-carnitine, lactate, pyruvate, ammonia, blood amino acid | CII + III↓ (fibroblasts) | dose not described, improvement of ICARS scoring after 3 months, the patient appeared to have a quicker response rate during conversation and better alertness *[Obj.]* | 17 years of age | Yair Anikster, Bruria Ben-Zeev, Edmond and Lily Safra Children's Hospital, Israel | (Malicdan et al., 2018) |

| | | | | | | | | | |
|---|---|---|---|---|---|---|---|---|---|
| | | | | | | profile, urine for protein and organic acids, were within normal limits. | | | |
| **COQ5 [2]** | | | ~ 66% (leukocytes) | childhood (F) | mild static gait ataxia, mild dysarthria, mild dysmetria and oculomotor apraxia, and horizontal nystagmus | no data | no data | dose not described, improvement of ICARS scoring after 3 months, the patient appeared to have a quicker response rate during conversation and better alertness *[Obj.]* | 22 years of age |
| **COQ5 [3]** | | | ~ 60% (leukocytes) | childhood (F) | mild motor delay, mild learning difficulties, mild cerebellar ataxia, mild cerebellar dysarthria and horizontal nystagmus | no data | no data | dose not described, improvement of ICARS scoring after 3 months, the patient appeared to have a quicker response rate during conversation and better alertness *[Obj.]* | 14 years of age |

**S1.6 Primary $CoQ_{10}$ deficiency-6 (COQ10D6; 614650) due to mutations in the *COQ6* gene [# of patients: 28]**

| Gene [Patient ID] | # of patients (# of families) | Mutation | Level of $CoQ_{10}$ (% of control)[1] | Age at onset (sex) if known | Symptoms | Biochemical tests and muscle pathology | RCC enzymes | $CoQ_{10}$ dose and response | Age at last reported exam or death | Corresponding PI | Reference |
|---|---|---|---|---|---|---|---|---|---|---|---|
| **COQ6 [1]** | 4 (1) | G255R (HOM) | no data | 6.4 y/o | SRNS, SND | no data | no data | not treated | 6.5 years of age; ESRF at age of 9.3 years | Friedhelm Hildebrandt, University of Michigan, USA | (Heeringa et al., 2011) |
| **COQ6 [2]** | | G255R (HOM) | no data | 0.3 y/o | SRNS, SND | no data | no data | not treated | died at age of 17.5 years | | |
| **COQ6 [3]** | | G255R (HOM) | no data | 1.2 y/o | SRNS, SND, ataxia | no data | no data | not treated | died at age of 6.5 years | | |
| **COQ6 [4]** | | not tested | no data | <1 y/o | SRNS, congenital SND | no data | no data | not treated | died at 5 years old | | |
| **COQ6 [5]** | 3 (1) | G255R (HOM) | no data | 0.3 y/o | SRNS, seizure | no data | no data | not treated | died, age of death not described | | |
| **COQ6 [6]** | | G255R (HOM) | no data | 0.3 y/o | SRNS, SND, facial dysmorphism | no data | no data | 100mg/day, improvement of SND *[NR]* | ESRF at age of 0.4 year | | |
| **COQ6 [7]** | | G255R (HOM) | no data | 0.2 y/o | SRNS, SND, bilateral nephrolithiasis | no data | no data | △30mg/kg/day beginning at 2 months of age (together with enalapril), a decrease of proteinuria, SND and severe growth retardation were noted at 10 months of age | 15 months of age | | |

| | | | | | | | | | | | |
|---|---|---|---|---|---|---|---|---|---|---|---|
| **COQ6 [8]** | 2 (1) | A353D (HOM) | no data | 6.0 y/o | SRNS, SND | no data | no data | not treated | ESFR at age of 6.5 years | | |
| **COQ6 [9]** | | A353D (HOM) | no data | 2.5 y/o | SRNS, SND | no data | no data | beginning at age 5.5 years, dose not described, decrease of proteinuria but no hearing improvement, reoccurrence of proteinuria after temporary cessation of $CoQ_{10}$ treatment and it decreased again after the treatment resumed *[Obj.]* | 6 years of age | | |
| **COQ6 [10]** | 1 (1) | A353D (HOM) | no data | 2.5 y/o | SRNS, seizure, white matter abnormalities | no data | no data | not treated | died, age of death unknown | | |
| **COQ6 [11]** | 1 (1) | W447X/ Q461fsX478 (CH) | no data | 3.0 y/o | SRNS, SND | no data | no data | not treated | 3 years of age | | |
| **COQ6 [12]** | 1 (1) | R162X/ ? | no data | no data | cyclosporine A-dependent NS | no data | no data | no data | no data | | |
| **COQ6 [13]** | 1 (1) | W188X/ ? | no data | no data | diffuse mesangial sclerosis | no data | no data | no data | no data | | |
| **COQ6 [14]** | 1 (1) | A353D (HOM) | no data | 4 y/o (M) | SRNS | no data | no data | no data | no data | Friedhelm Hildebrandt, Boston Children’s Hospital, USA | (Sadowski et al., 2015) |
| **COQ6 [15]** | 1 (1) | A353D (HOM) | no data | 3.2 y/o (M) | SRNS | no data | no data | no data | no data | | |
| **COQ6 [16]** | 1 (1) | D385A/ Y412C (CH) | no data | 4.5 y/o (M) | SRNS | no data | no data | no data | no data | | |
| **COQ6 [17]** | 1 (1) | R360W/ c.804delC (CH) | no data | 2 y/o (F) | steroid-resistant glomerulopathy, poor growth | proteinuria (mostly during respiratory tract infection) | no data | 30 mg/kg/day, remission of glomerulopathy after 1 month of treatment, growth acceleration after 12 months and a reduction of respiratory airway infections *[NR]* | 4 years of age | Małgorzata Stańczyk University of Lodz, Poland | (Koyun et al., 2019; Stanczyk et al., 2018) |
| **COQ6 [18]** | 1 (1) | P261L (HOM) | no data | 0.8 y/o (M) | SRNS | no data | no data | treated, response not described | 4 years of age | L Gesualdo, University of Bari Aldo Moro, Italy | (Gigante et al., 2017) |
| **COQ6 [19]** | 1 (1) | K64del/ P261L (CH) | no data | 3.8 y/o (M) | steroid-resistant FSGS, mild muscle weakness in the lower extremities | no data | no data | no data | no data | Hae Il Cheong, Seoul National University Hospital, South Korea | (Park et al., 2017a) |
| **COQ6 [20]** | 1 (1) | K64del/ Q229P (CH) | no data | 1.8 y/o (F) | SR-FSGS, exotropia with nystagmus | no data | no data | no data | no data | | |
| **COQ6 [21]** | 1 (1) | K64del/ P261L (CH) | no data | 3.9 y/o (F) | SR-FSGS | no data | no data | no data | no data | | |
| **COQ6 [22]** | 1 (1) | K64del/ P261L | no data | 2.7 y/o (F) | SR-FSGS | no data | no data | no data | no data | | |

| | | | | | | | | | | | |
|---|---|---|---|---|---|---|---|---|---|---|---|
| | | (CH) | | | | | | | | | |
| **COQ6 [23]** | 1 (1) | K64del/ P261L (CH) | no data | 1.3 y/o (F) | SR-FSGS, optic nerve atrophy | no data | no data | no data | no data | | |
| **COQ6 [24]** | 1 (1) | K64del/ P261L (CH) | no data | 2.1 y/o (M) | SR-FSGS, mild muscle weakness in the lower extremities | no data | no data | no data | no data | | |
| **COQ6 [25]** | 2 (1) | A353D (HOM) | no data | 5 y/o (M) | SRNS, SND, optic atrophy | no data | normal CI+CIII and CII+CIII activities (muscle) | △15mg/kg/day of idebenone beginning at age of 17 years after the onset of optical symptoms, an improvement in the visual acuity after 2 months of treatment. After 13 months of treatment, the optical examination was stable, but the patient did not recover normal vision, still exhibiting persistent optic atrophy. After 3 years of treatment, minimal optic atrophy was reported. No change of the deafness status since treatment initiation. [other medications: immunosuppressive treatment] | Age of 18 years, kidney transplant at age 6 | Justine Perrin, Hôpital Sainte-Musse, France | (Justine Perrin et al., 2020) |
| **COQ6 [26]** | | A353D (HOM) | no data | 4 y/o (M) | SRNS, SND | no data | no data | △10mg/kg/day of idebenone since age 7, after 13 months of treatment, hearing loss was not changed and renal involvement remained stable with only Enalapril, demonstrated by negative proteinuria. | 8 years of age | | |
| **COQ6 [27]** | 2 (1) | Y83X/Q 461 (CH) | no data | 4 m/o (F) | seizure, growth retardation, proteinuria, atrial septal defect, and pulmonary hypertension | blood lactate↑, lipids ↑, albumin↓, urine organic acid ↑ | no data | not treated | died at age of 5 months | Lizhen Wang, Wenzhou Medical University | (Wang et al., 2021) |
| **COQ6 [28]** | | | no data | 3 m/o (M) | proteinuria, growth retardation, and muscle hypotonia | blood lactate↑, triglyceride ↑, albumin↓, edema | no data | not treated | died at age of 4 months | | |

**S1.7 Primary CoQ$_{10}$ deficiency-8 (COQ10D8; 616733) due to mutations in the *COQ7* gene [# of patients: 6]**

| Gene [Patient ID] | # of patients (# of families) | Mutation | Level of CoQ$_{10}$ (% of control)[1] | Age at onset (sex) if known | Symptoms | Biochemical tests and muscle pathology | RCC enzymes | CoQ$_{10}$ dose and response | Age at last reported exam or death | Corresponding PI | References |
|---|---|---|---|---|---|---|---|---|---|---|---|

| | | | | | | | | | | | |
|---|---|---|---|---|---|---|---|---|---|---|---|
| **COQ7 [1]** | 1 (1) | V141E (HOM) | ~ 10% (fibroblasts, muscle) | neonatal (M) | muscular hypotonia, developmental retardation, learning disabilities, hearing impairment, visual dysfunction, not able to sit and walk independently | blood and CSF lactate↑/small fiber size, no abnormal mitochondrial structure observed in the muscle | CI+CIII ↓, CII+CIII ↓ (fibroblasts), CI+CIII ↓, CI ↓ (muscle) | initially treated with idebenone, switched to $COQ_{10}$ after the diagnosis of a primary $CoQ_{10}$ deficiency (around age of 10 years), dosage unknown, stalling the regression and significantly reducing the pain were noted *[NR]* | 9 years of age | Anna Wredenberg, Karolinska Institutet, Sweden | (Freyer et al., 2015) |
| **COQ7 [2]** | 1 (1) | L111P (HOM) | ~ 70% (fibroblasts) | 14 m/o (F) | spasticity, muscle wasting, inability to walk without support | CSF lactate↑ | no data | 22.8 mg/kg/day, no response after 3 months of treatment *[NR]* | 6 years of age | Siegfried Hekimi, McGill University, Canada | (Wang et al., 2017b) |
| **COQ7 [3]** | 1 (1) | K200Ifs X56/ R107W (CH) | ~ 12% (fibroblasts) | neonatal (M) | cardiomyopathy, growth retardation, hypotonia, ptosis, visual impairment, hearing impairment, muscle weakness, infantile spasms | blood lactate and alanine↑, urinary lactate↑, pyruvate↑, and 3-hydroxybutyrate↑, dicarboxylic aciduria, excretions of Kreb cycle intermediates↑ | no data | Beginning at 2 months of age, and the dose was increased to 20 mg/kg/day at 12 months of life, the patient died around the same time *[NR]* | 1 year of age | Cheuk-Wing Fung and Brian H.-Y. Chung, Queen Mary Hospital, China | (Kwong et al, 2019) |
| **COQ7 [4]** | 1 (1) | R54Q (HOM) | ~ 55 % (fibroblasts) | 15 m/o (M) | hypotonia, difficulty walking, motor developmental delay, ataxia, and spasticity | no data | no data | not treated | 6 years of age | Evren Gumus, Mugla Sitki Kocman University, Turkey | https://doi.org/10.1016/j.ymgmr.2022.100877 |
| **COQ7 [5]**<br>**COQ7 [6]** | 2 (1) | I66N/Y149C (CH) | no data | Pediatric (unknown) | axonal neuropathy, mild neurodegenerative disorder | no data | no data | no data | no data | Hubert Smeets, Maastricht University, Netherlands | (Theunissen et al., 2018) |

#### S1.8 Primary $CoQ_{10}$ deficiency-4 (COQ10D4; 612016) due to mutations in the *COQ8A/ADCK3* gene [# of patients: 112]

| Gene [Patient ID] | # of patients (# of families) | Mutation | Level of $CoQ_{10}$ (% of control)[1] | Age at onset (sex) if known | Symptoms | Biochemical tests and muscle pathology | RCC enzymes | $CoQ_{10}$ dose and response[2] | Age at last reported exam or death | Corresponding PI | References |
|---|---|---|---|---|---|---|---|---|---|---|---|
| **COQ8A [1]** | 2 (1) | R213W/ G272V (CH) | ~ 29% (muscle) | 18 m/o (F) | hypotonia, *talus valgus*, developmental delay, seizure, ataxia, epilepsia partialis continua | blood and CSF lactate in normal range | CI↑, CII↑, CIII↑, CIV↑ (muscle) | 20 mg/kg/day (350mg/day) for 8 years, no response *[NR]* | 21 years of age | Agnès Rötig, Hôpital Necker-Enfants Malades, France | (Mignot et al., 2013; Mollet et al., 2008) |
| **COQ8A [2]** | | R213W/ G272V (CH) | no data | 2 y/o (F) | hypotonia, seizure, ataxia, developmental delay | blood and CSF lactate in normal range | no data | 350mg/day for 13 months, no response *[NR]* | 15 years of age | | |
| **COQ8A [3]** | 1 (1) | E551K (HOM) | ~ 8% | 18 m/o (M) | cerebella ataxia, strabismus, muscle weakness, trunk hypotonia, tonic seizure | blood lactate↑, no | CI+CIII↓(muscle) | 5mg/kg/day from age 3 years, 10mg/kg/day from age 4 to 7, no response; followed by | 16 years of age | | (Mollet et al., 2008) |

| | | | | | | | | | | | |
|---|---|---|---|---|---|---|---|---|---|---|---|
| | | | (muscle), normal range (fibroblasts) | | | ragged-red fibers in the muscle but mitochondrial accumulation and lipid droplets in 10%–20% of the fibers | | 10mg/kg/day of idebenone for 7 months which worsened the patient's conditions *[NR]* | | | |
| **COQ8A [4]** | 1 (1) | G272D/ Q605GfsX 125 (CH) | < 5% (muscle), normal range (fibroblasts) | 3 y/o (F) | exercise intolerance, muscle weakness, cerebellar syndromes, seizure | blood lactate↑, mitochondrial myopathy in the muscle | CI↑, CII↑, CIII↑, CIV↑, CS↑, CI+CIII↓, CII+CIII↓ (muscle) | △6 mg/kg/day (750mg/day) of $CoQ_{10}$ and L-carnitine were initiated at age 5, improved exercise tolerance and fewer vomiting episodes were noted after 3 months of therapy. $CoQ_{10}$ was replaced with idebenone (5mg/kg/day) at the age of 9 years, and within the following 4 months, severe exercise intolerance reappeared with numerous episodes of vomiting. Reverting to $CoQ_{10}$ treatment resulted in returns to the previous clinical status within 3 months. *[Obj.]* | 20 years of age | Anne Lombès, Hospitalier Pitié-Salpêtrière, France | (Aure et al., 2004; Mignot et al., 2013; Mollet et al., 2008) |
| **COQ8A [5]** | 4 (1) | c.1398+2T →C (D420WfsX 40, I67AfsX22) (HOM) | no data | 11 y/o (M) | cerebellar ataxia | no data | no data | not treated | 42 years of age | Michel Koenig, Hôpitaux Universitaires de Strasbourg, France | (Lagier-Tourenne et al., 2008; Quinzii et al., 2010) |
| **COQ8A [6]** | | c.1398+2T →C (D420WfsX 40, I67AfsX22) (HOM) | no data | 4 y/o (M) | cerebellar ataxia, exercise intolerance | blood lactate↑ | no data | not treated | 38 years of age | | (Lagier-Tourenne et al., 2008) |
| **COQ8A [7]** | | c.1398+2T →C (D420WfsX 40, I67AfsX22) (HOM) | normal range (fibroblasts) | 7 y/o (M) | Cerebellar ataxia, exercise intolerance | blood lactate↑ | CI+CIII↓ (fibroblasts) | not treated | 36 years of age | | (Lagier-Tourenne et al., 2008) |
| **COQ8A [8]** | | c.1398+2T →C (D420WfsX 40, I67AfsX22) (HOM) | no data | 8 y/o (F) | Cerebellar ataxia, exercise intolerance | blood lactate↑ | no data | not treated | 29 years of age | | (Lagier-Tourenne et al., 2008) |
| **COQ8A [9]** | 1 (1) | Q167LfsX 36 | ~ 64% (fibroblasts) | 4 y/o (M) | cerebellar ataxia, mild mental retardation | blood lactate in normal range | CI+CIII ↓ , CII+CIII ↓ (fibroblasts) | not treated | 18 years of age | | (Lagier-Tourenne |

| | | | | | | | | | | | |
|---|---|---|---|---|---|---|---|---|---|---|---|
| | | (HOM) | | | | | | | | | et al., 2008; Quinzii et al., 2010) |
| **COQ8A [10]** | 1 (1) | Y514C/ T584del (CH) | ~ 51% (fibroblasts), ~ 46% (muscle) | 5 y/o (M) | cerebellar ataxia, gynecomastia, feet and thumbs in dystonic position | blood lactate in normal range | CI+CIII ↓ , CII+CIII ↓ (fibroblasts) | 60 -700 mg/day over 8 years, the patient reported mild subjective improvement, and stabilization of the cerebellar ataxia was observed on examination | 17 years of age | | (Lagier-Tourenne et al., 2008; Lamperti et al., 2003; Quinzii et al., 2010) |
| **COQ8A [11]** | 1 (1) | K314_Q360 del/ G549S (CH) | no data | 3 y/o (F) | cerebellar ataxia, mild hearing loss | blood lactate in normal range | no data | not treated | 30 years of age | | (Lagier-Tourenne et al., 2008) |
| **COQ8A [12]** | 3 (1) | R348X (HOM) | no data | 3 y/o (M) | cerebellar ataxia, exercise intolerance, myoclonus, tremor, dystonic posture | no data | no data | no data | 31 years of age | Hubert Smeets, Maastricht University, The Netherlands | (Gerards et al., 2010) |
| **COQ8A [13]** | | R348X (HOM) | no data | 9 y/o (M) | cerebellar ataxia, cognitive impairment, speech and coordination difficulties, exercise intolerance | no data | no data | no data | 26 years of age | | |
| **COQ8A [14]** | | R348X (HOM) | no data | 3 y/o (M) | cerebellar ataxia, epilepsy, exercise intolerance, vision impairment | no data | no data | no data | 25 years of age | | |
| **COQ8A [15]** | 2 (1) | R348X/ L379X (CH) | no data | 2 y/o (F) | ataxia, tremor, dysarthric and monotonous speech, exercise intolerance, slight spasticity | unremarkable muscle morphology | no data | no data | 26 years of age | | |
| **COQ8A [16]** | | R348X/ L379X (CH) | no data | infancy (M) | cerebellar ataxia, dysarthria | unremarkable muscle morphology | CII+CIII↓ (muscle) | no data | 21 years of age | | |
| **COQ8A [17]** | 1 (1) | R348X (HOM) | <14.5% (muscle) | 6 y/o (F) | seizure, ataxia, cerebellar atrophy, a mild cognitive delay | laboratory tests, including creatine kinase, and metabolic investigations, including transferrin isofocusing, serum lactate, serum and urine organic acids, were unremarkable | CII+CIII ↓ (muscle) | 10mg/kg/day initiated at the age of 8 years, within 6 months improvement of ataxia was observed, but after 5 years of treatment, MRI showed increased cerebellar atrophy | 17 years of age | Enrico Bertini, Bambino Gesù Children's Hospital, Italy | (Terracciano et al., 2012) |
| **COQ8A [18]** | 2 (1) | T584delA CC/ P502R (CH) | no data | 2 y/o (F) | cerebellar ataxia, dysarthria, nystagmus, cognitive decline, psychiatric disorder | metabolic evaluation and muscle | CI+CIII↓, CIV↓(muscle) | 20mg/kg/day initiated at age 5, partial improvement in motor skills, balance, and strength; after 6 years, | 20 years of age | Dorit Lev, Wolfson Medical Center, Israel | (Blumkin et al., 2014) |

| | | | | | | | | | | | |
|---|---|---|---|---|---|---|---|---|---|---|---|
| | | | | | | morphology were unremarkable | | treatment was discontinued, and the patient’s condition deteriorated. *[Obj.]* | | | |
| **COQ8A [19]** | | T584delACC/ P502R (CH) | no data | childhood (F) | mild dysfluent speech and clumsiness, cerebellar atrophy, mild dysarthria | no data | no data | treated, dosage and response not described | 32 years of age | | |
| **COQ8A [20]** | 1 (1) | S616LfsX 114/ R301Q (CH) | ~ 45% (plasma) | 9 y/o (M) | exercise intolerance, cerebellar ataxia, tremors, dysautonomia | blood lactate↑, the remaining blood tests, including liver function and serum creatine kinase were all normal | no data | 120mg/day, self-reported fatigue and exercise tolerance improved after 2 weeks of therapy. After 2 years of therapy, ataxia and head tremor diminished and SARA total score improved. When the treatment was stopped for a month, the patient’s condition deteriorated, rendering him to resume taking $CoQ_{10}$. *[Obj.]* | 35 years of age | Dantao Peng, China-Japan Friendship Hospital, China | (Zhang et al., 2020) |
| **COQ8A [21]** | 1 (1) | R271C/ A304T (CH) | normal range (muscle) | 15 y/o (F) | cerebellar ataxia, tremors | no data | COX↓(muscle) | 300 mg/day, no response after 6 months *[NR]* | 46 years of age | Rita Horvath, Newcastle University, UK | (Horvath et al., 2012) |
| **COQ8A [22]** | 1 (1) | A304V (HOM) | ~ 8% (muscle) | 27 y/o (F) | cerebellar ataxia, upper-limb myoclonus, seizure, dysmetria, cataract | no data | CI↓, CIV↓, COX↓, lipid↑ (muscle) | 300 mg/day, no response after 6 months *[NR]* | 50 years of age | | |
| **COQ8A [23]** | 1 (1) | R299W (HOM) | no data | 1 y/o (F) | cerebellar ataxia, seizure, mental retardation, unable to walk by 12 years | no data | no data | 200 mg/day, no response within 2 months *[NR]* | 18 years of age | | |
| **COQ8A [24]** | 1 (1) | Y429C/? | ~ 22% (muscle) | 1.5-2 y/o (F) | ataxia, muscle weakness, cognitive impairment, horizontal nystagmus, bilateral dysmetria, tremors | no data | CI↓, CIV↓, CII+CIII↓, COX↓, lipid ↑ (muscle) | 200 mg/day, no response within 2 months *[NR]* | 20 years of age | | |
| **COQ8A [25]** | 2 (1) | S616LfsX 114 (HOM) | ~ 35% (fibroblasts) | 10 y/o (F) | cerebellar ataxia, myoclonus, slurred speech, wheelchair-dependent by 30 years of age | no data | CI↓, CII+CIII↓ (fibroblasts) | 400mg/day, improvement in myoclonic symptoms, speech quality (after 3 months), and ataxia with a reduction in SARA (after 6 months) *[Obj.]* | 35 years of age | Henry Houlden, National Hospital for Neurology and Neurosurgery, UK | (Liu et al., 2014) |
| **COQ8A [26]** | | S616LfsX 114 (HOM) | no data | 14 y/o (M) | cerebellar ataxia, myoclonus, tremors, dysarthric speech | no data | no data | 200mg/day, improvement in speech and fatigue after 3 months of treatment | 32 years of age | | |
| **COQ8A [27]** | 1 (1) | R301W/ c.1399-3_ 1408del (CH) | low (muscle) | 11 y/o (M) | reduced dexterity, dysarthria, hypometric saccades, scanning speech, and dystonic posturing, tremors, ataxia | no data | no data | 800mg/day, a resolution of tremors and improvement of limb and truncal dystonia after 9 months of treatment *[Subj.]* | 25 years of age | Renato Puppi Munhoz, University of Toronto, Canada | (Chang et al., 2018) |
| **COQ8A [28]** | 1 (1) | T584del/ T511M (CH) | low (muscle) | 10 y/o (F) | ataxia, tremors, dysarthria, appendicular dysmetria, truncal instability, titubation, wheelchair-dependent by 53 years of age | no data | no data | 800mg/day, improvement of ataxia overall with a reduction in SARA score, able to work independently, | 54 years of age | | |

| | | | | | | | | | | | |
|---|---|---|---|---|---|---|---|---|---|---|---|
| | | | | | | | | after 9 months of therapy. *[Obj.]* | | | |
| **COQ8A [29]** | 1 (1) | D305Y (HOM) | low (muscle) | 5 y/o (M) | developmental delay, intellectual disability, ataxia, isolated pan-cerebellar features including head titubation, dysmetria, dysdiadochokinesia | no data | normal range of activities of CI and CII, CS and COX (muscle) | 800mg/day, inconsistent use for 2 years, no response *[NR]* | 33 years of age | | |
| **COQ8A [30]** | 1 (1) | T445RfsX 52 (HOM) | no data | no data (F) | ataxia, seizure, developmental delay, strabismus | no data | no data | no data | 15 years of age | Miao Sun, The University of Chicago, USA | (Sun et al., 2019) |
| **COQ8A [31]** | 1 (1) | T511M (HOM) | no data | no data (F) | ataxia, developmental delay | no data | no data | no data | 20 years of age | | |
| **COQ8A [32]** | 1 (1) | R348X/ 2A>G [p?] (CH) | no data | no data (F) | ataxia, dysarthria | no data | no data | no data | 45 years of age | | |
| **COQ8A [33]** | 1 (1) | R271C/ R334W (CH) | no data | 25 y/o (M) | ataxia, dystonia, myoclonus, tremors, seizure | no data | no data | no data | 31 years of age | | |
| **COQ8A [34]** | 1 (1) | E551K/ R301W (CH) | no data | no data (F) | ataxia, seizure | no data | no data | no data | 33 years of age | | |
| **COQ8A [35]** | 1 (1) | R301W/ R410Q (CH) | no data | no data (F) | ataxia, developmental delay | no data | no data | no data | 8 years of age | | |
| **COQ8A [36]** | 1 (1) | N148X/ A338T (CH) | no data | 12 y/o (F) | cerebellar ataxia, tremors, focal dystonia | blood lactate in normal range | no data | not treated | 35 years of age | Matthis Synofzik University of Tübingen, Germany | (Traschutz et al., 2020) |
| **COQ8A [37]** | 1 (1) | A42fs/ Q50X (CH) | no data | no data (F) | cerebellar ataxia, dystonic tremor | no data | no data | not treated | 38 years of age | | |
| **COQ8A [38]** | 1 (1) | A339T/ Y361 (CH) | no data | 42 y/o (M) | cerebellar ataxia, stroke-like episode, muscle weakness, hearing loss | blood lactate in normal range, no ragged red fibers in the muscle | activities in normal range | dosage not described, no response *[NR]* | 45 years of age | | |
| **COQ8A [39]** | 2 (1) | A337T (HOM) | no data | 6 y/o (M) | cerebellar ataxia, dystonia, tremor, | blood lactate in normal range | no data | 600mg/day, no response *[NR]* | 12 years of age | | |
| **COQ8A [40]** | | A337T (HOM) | no data | 2 y/o (M) | ataxia, impairment of speech | no data | no data | not treated | 6 years of age | | |
| **COQ8A [41]** | 1 (1) | A338V (HOM) | no data | 13 y/o (F) | cerebellar ataxia, muscle weakness, myoclonus, tremor, dysarthria | blood lactate in normal range | no data | dosage not described, improved tremors | 18 years of age | | |
| **COQ8A [42]** | 2 (1) | V83fs (HOM) | no data | 8 y/o (F) | cerebellar ataxia, tremors | blood lactate ↑ | no data | 1250mg/day, improved tremors | 37 years of age | | |
| **COQ8A [43]** | | V83fs (HOM) | no data | 16 y/o (M) | cerebellar ataxia, dysarthria, tremors | blood lactate in normal range | no data | 1250mg/day, response not described | 25 years of age | | |
| **COQ8A [44]** | 1 (1) | T584del/ A338T (CH) | ~ 15% (muscle) | 6 y/o (F) | ataxia, pan-cerebellar atrophy | mild mitochondrial myopathy with ragged red fibers | normal RC enzyme activities (muscle) | 100mg/day, response not described | 69 years of age | | |

| | | | | | | | | | |
|---|---|---|---|---|---|---|---|---|---|
| | | | | | | and COX- fibers (muscle) | | | |
| **COQ8A [45]** | 1 (1) | E481X (HOM) | no data | 1 y/o (F) | ataxia, motor retardation, cognitive impairment, tremors | blood lactate in normal range | no data | 200mg/day, no initial apparent effect but after stop: fatigue and falls; improvement of muscle weakness with reintroduction of $CoQ_{10}$ | 16 years of age |
| **COQ8A [46]** | 1 (1) | Q167LfsX 36/ R348X (CH) | no data | 1 y/o (M) | ataxia | no data | no data | not treated | 17 years of age |
| **COQ8A [47]** | 1 (1) | R271C/ T487R (CH) | no data | 6 y/o (F) | ataxia, seizure, stroke-like episodes | unremarkable muscle histology | normal range (muscle) | not treated | 26 years of age |
| **COQ8A [48]** | 1 (1) | c.589-3C>G/ G615D (CH) | no data | 2 y/o (F) | ataxia, hypotonia | blood lactate in normal range | no data | 10 mg/kg/day, improvement in stability | 9 years of age |
| **COQ8A [49]** | 1 (1) | R348X (HOM) | no data | 25 y/o (M) | ataxia, tremors, spasticity | no data | no data | not treated | 53 years of age |
| **COQ8A [50]** | 1 (1) | c.589-3C>G/ R301W (CH) | no data | 2 y/o (F) | ataxia, seizure | no data | no data | 10 mg/kg/day, improved balance | 13 years of age |
| **COQ8A [51]** | 2 (1) | R301W/ E446AfsX 33 (CH) | low (muscle) | 3 y/o (M) | ataxia | mild muscle histology changes | mild changes (muscle) | 10 mg/kg/day, no response *[NR]* | 10 years of age |
| **COQ8A [52]** | | R301W/ E446AfsX 33 (CH) | no data | 2 y/o (M) | ataxia, developmental retardation | no data | no data | 10 mg/kg/day, no response *[NR]* | 7 years of age |
| **COQ8A [53]** | 1 (1) | R348X (HOM) | low (muscle) | 10 y/o (F) | epilepsy, ataxia | blood lactate in normal range | normal range (muscle) | 600mg/day, no response *[NR]* | 24 years of age |
| **COQ8A [54]** | 1 (1) | R301W (HOM) | low (muscle) | 8 y/o (F) | ataxia, seizure, cardiomyopathy | blood lactate in normal range | normal range (muscle) | 400mg/day, no response *[NR]* | death at 17 years of age |
| **COQ8A [55]** | 1 (1) | E568X (HOM) | no data | 6 y/o (F) | spastic hypertonia, ataxia | no data | no data | 300mg/day since 5 years old, more energetic, mentally quicker | 69 years of age |
| **COQ8A [56]** | 2 (1) | M555I (HOM) | no data | 11 y/o (F) | ataxia, exercise intolerance, cognitive complaints | no data | no data | not treated | 46 years of age |
| **COQ8A [57]** | | M555I (HOM) | no data | 1 y/o (F) | ataxia, memory/concentration difficulties | unremarkable muscle histology | no data | not treated | 40 years of age |
| **COQ8A [58]** | 1 (1) | Q167Lfs (HOM) | no data | 1 y/o (M) | developmental delay, hypomimia, learning difficulties, ataxia | blood lactate↑ | no data | not treated | 19 years of age |
| **COQ8A [59]** | 1 (1) | O207L (HOM) | no data | 11 y/o (F) | ataxia, tremors, myoclonus | no data | no data | not treated | 19 years of age |

| | | | | | | | | | | | |
|---|---|---|---|---|---|---|---|---|---|---|---|
| **COQ8A [60]** | 1 (1) | H85AfsX42 (HOM) | no data | 3 y/o (M) | ataxia | blood lactate in normal range | no data | 400 - 1200 mg/day, response not described | 9 years of age | | |
| **COQ8A [61]** | 1 (1) | L275RfsX16/ L402P (CH) | no data | 6 y/o (F) | ataxia, choreiform dyskinesia, tremors | blood lactate in normal range | no data | not treated | 16 years of age | | |
| **COQ8A [62]** | 2 (1) | C268R (HOM) | no data | 6 y/o (F) | ataxia, epilepsy | no data | no data | not treated | 23 years of age | | |
| **COQ8A [63]** | | C268R (HOM) | no data | 2 y/o (F) | developmental delay, epilepsy, cerebellar atrophy | no data | no data | not treated | 21 years of age | | |
| **COQ8A [64]** | 1 (1) | M555I (HOM) | no data | 1 y/o (M) | hearing loss, tremors, cerebellar atrophy | no data | no data | not treated | 59 years of age | | |
| **COQ8A [65]** | 1 (1) | R301W/ M555I (CH) | no data | 4 y/o (F) | mental and motor retardation, ataxia, tremors, seizure | blood lactate in normal range | no data | not treated | 58 years of age | | |
| **COQ8A [66]** | 1 (1) | G342W (HOM) | no data | 10 y/o (M) | ataxia, tremors | no data | no data | not treated | 78 years of age | | |
| **COQ8A [67]** | 2 (1) | R213G (HOM) | no data | childhood (M) | ataxia, speech and swallowing difficulties, leg cramps | no data | no data | not treated | 63 years of age | | |
| **COQ8A [68]** | | R213G (HOM) | no data | 20 y/o (F) | ataxia, seizure, speech and swallowing difficulties | blood lactate in normal range | no data | not treated | 58 years of age | | |
| **COQ8A [69]** | 1 (1) | I4K/ R512W (CH) | no data | 2 y/o (M) | mental retardation, ataxia | no data | no data | not treated | 18 years of age | | |
| **COQ8A [70]** | 1 (1) | L453RfsX24/ E568X (CH) | no data | 13 y/o (F) | tremors, incoordination | blood lactate in normal range | no data | not treated | 37 years of age | | |
| **COQ8A [71]** | 1 (1) | L453RfsX24/ E568X (CH) | no data | 22 y/o (F) | ataxia, bipolar disorder, impulsive behavior | no data | no data | not treated | 71 years of age | | |
| **COQ8A [72]** | 2 (1) | T445fs (HOM) | no data | 7 y/o (F) | mild ataxia, mild dysarthria | no data | no data | not treated | 41 years of age | | |
| **COQ8A [73]** | | T445fs (HOM) | no data | 6 y/o (M) | mild ataxia, mild dysarthria tremors, speech disorder | mild denervation in muscle | no data | not treated | 37 years of age | | |
| **COQ8A [74]** | 2 (1) | G615D (HOM) | no data | childhood (M) | ataxia, dysmetria, seizure | blood lactate in normal range | no data | 135mg/day of idebenone for 9 months, response not described | 29 years of age | | |
| **COQ8A [75]** | | G615D (HOM) | no data | 7 y/o (F) | ataxia. dysmetria | no data | no data | 135mg/day of idebenone, response not described | 24 years of age | | |
| **COQ8A [76]** | 1 (1) | del exons 3-15/ F508S (CH) | no data | 6 y/o (M) | ataxia. dysmetria, myoclonus | no data | no data | 300mg/day for 15 months, improvement in movement disorder and SARA score *[Obj.]* | 17 years of age | Mathieu Anheim, Hôpital de la Salpêtrière, France | (Mignot et al., 2013) |
| **COQ8A [77]** | 1 (1) | R299W/ L453RfsX24 | normal range (fibroblasts) | 15 y/o (M) | ataxia, seizure, myoclonus, dysmetria | no data | no data | 300mg/day for 8 months, improvement in movement disorder *[Subj.]* | 44 years of age | | |

| | | | | | | | | | | | |
|---|---|---|---|---|---|---|---|---|---|---|---|
| | | (CH) | | | | | | | | | |
| **COQ8A [78]** | 2 (1) | R299W/ R410X (CH) | no data | 4 y/o (F) | ataxia, dysmetria, seizure | no data | no data | 300mg/day for1 month, withdrawn, reversible side effect of treatment (anorexia) *[NR]* | 38 years of age | | |
| **COQ8A [79]** | | R299W/ R410X (CH) | no data | 4 y/o (M) | ataxia, dysmetria, seizure | blood lactate in normal range | no data | 300mg/day for1 month, withdrawn, reversible side effect of treatment (diarrhea) *[NR]* | 34 years of age | | |
| **COQ8A [80]** | 1 (1) | R271C (HOM) | low (plasma) | 1.5 y/o (F) | ataxia, seizure, dystonia, chorea, dysmetria, myoclonus, spasticity | blood lactate in normal range | CII+CIII↓ (muscle) | 30 mg/kg/day for 3 years, no response *[NR]* | 5 years of age | | |
| **COQ8A [81]** | 2 (1) | L197VfsX 20 (HOM) | no data | 19 y/o (F) | ataxia. dysmetria | blood lactate in normal range | no data | 1200mg/day for 1 year no response *[NR]* | 34 years of age | | |
| **COQ8A [82]** | | L197VfsX 20 (HOM) | no data | 19 y/o (F) | ataxia. dysmetria, seizure | blood lactate in normal range | no data | 1200mg/day for 1 year, no response *[NR]* | 31 years of age | | |
| **COQ8A [83]** | 1 (1) | Q360_Y36 1insX (HOM) | no data | 2 y/o (F) | ataxia. Dysmetria, tremors | blood lactate in normal range | no data | 800mg/day for 1 year, no response *[NR]* | 15 years of age | | |
| **COQ8A [84]** | 1 (1) | R299W (HOM) | ~ 10-24% (muscle) | 7 y/o (F) | ataxia, seizure, tremor | unremarkable muscle biopsy | no data | 900mg/day for 6 months, no response *[NR]* | 35 years of age | L. A. Bindof, University of Bergen, Norway | (Hikmat et al., 2016) |
| **COQ8A [85]** | 2 (1) | R299W/ F578V (CH) | ~ 34-60% (muscle) | 7 y/o (M) | ataxia, seizure, dysmetria, tremors, dysarthria, dysdiadochokinesia | unremarkable muscle biopsy | no data | 600mg/day since the age of 33, improvement in balance and coordination (reported by the patient) and a reduction of SARA score *[Obj.]* | 34 years of age | | |
| **COQ8A [86]** | | R299W/ F578V (CH) | no data | 3 y/o (F) | dysarthria, ataxia, epilepsy, cognitive impairment, tremors | no data | no data | no data | died at age of 22 | | |
| **COQ8A [87]** | 1 (1) | R299W (HOM) | no data | 2 y/o (F) | ataxia, epilepsy, seizure, feeding difficulties | unremarkable muscle biopsy | no data | 1000mg/day of deoxyubiquinone (probably ubiquinol) since age of 18, no response *[NR]* | 22 years of age | | |
| **COQ8A [88]** | 1 (1) | R299W/ E551K (CH) | no data | 5 y/o (M) | dysarthria, ataxia, seizure, delayed growth | blood lactate in normal range | no data | no data | 18 years of age | Mathieu Anheim, Hôpital de Hautepierre, France | (Mallaret et al., 2016) |
| **COQ8A [89]** | 1 (1) | A304V (HOM) | no data | 10 y/o (M) | mild developmental delay, ataxia | no data | no data | no data | 41 years of age | | |
| **COQ8A [90]** | 1 (1) | R299W/ L453RfsX 24 (CH) | no data | 15 y/o (M) | mild developmental delay, ataxia | no data | no data | no data | 46 years of age | | |
| **COQ8A [91]** | 1 (1) | 27.6 kb deletion of 1q42.3 involving exons 1 and 2 | ~ 34% (muscle), normal range (fibroblasts) | 13 y/o (F) | ataxia, tremors, hand bradykinesia, subtle and variable speech dysfluency | no data | CI+CIII↓, CII+CIII↓, CS ↓ (muscle) | △ Tremor improved on trihexyphenidyl/clonazepam combination therapy before ubiquinol supplementation which was initiated at age 19 years. Ubiquinol dosage was | 25 years of age | Jennifer Friedman, Rady Children's Hospital, USA | (Galosi et al., 2019) |

| | | | | | | | | | | | |
|---|---|---|---|---|---|---|---|---|---|---|---|
| | | (HOM) | | | | | | not described. After two years of ubiquinol and high-dose vitamin B-complex treatments, tremor was stable, and the patient was able to tandem walk normally. She had marked bradykinesia though. | | | |
| **COQ8A [92]** | 1 (1) | G615D/ L197VfsX 20 (CH) | no data | 7 y/o (F) | tremors, ataxia, dysmetria, difficulty writing and hand clumsiness | metabolic work-up, including, plasma amino acids, plasma acyl-carnitine profile, urinary organic acids, lactate, was normal | no data | 800mg/day initiated at the age of 8.5, clinical stabilization was reported after the treatment | 10 years of age | | |
| **COQ8A [93]** | 1 (1) | R348X (HOM) | no data | 25 y/o (M) | ataxia, tremors, writing difficulties | Blood creatine kinase and cholesterol ↑, the remaining blood biochemistry including urinary organic acids was normal | no data | no data | 54 years of age | | |
| **COQ8A [94]** | 1 (1) | R301W/ E446AfsX 33 (CH) | no data | 3 y/o (M) | ataxia, speech difficulties, seizure, tremors, dystonia | no data | no data | 10 mg/kg/day, initiated at age 10, but has been taken only intermittently, response not described | 11 years of age | | |
| **COQ8A [95]** | 2 (1) | L277P/ c.1506+1 G>A (CH) | low (muscle) normal range (plasma) | childhood (F) | ataxia, dysmetria, hypotonia | The metabolic workup, including plasma cholesterol, plasma albumin, plasma and urinary amino acids, urinary organic acids, and CSF lactate was normal. Mitochondria in the muscle were unremarkable. | CII+CIII↓ (muscle) | 20 mg/kg/day, improvement in an ataxia assessment score at 1-year follow-up *[Obj.]* | 7.8 years of age | R. G. Snell, The University of Auckland, New Zealand | (Jacobsen et al., 2018) |
| **COQ8A [96]** | | L277P/ c.1506+1 G>A (CH) | normal range (plasma) | childhood (F) | ataxia | | CII+CIII↓ (muscle) | 20 mg/kg/day, minimal improvement in an ataxia assessment score at 1-year follow-up *[NR]* | 2.2 years of age | | |
| **COQ8A [97]** | 1 (1) | c.655+1G >A/ A339T (CH) | no data | 3 y/o (F) | exercise intolerance, dysarthria, seizure, stroke-like episodes, ataxia, homonymous hemianopsia, dysarthria | blood creatinine kinase↑, blood lactate↑, CI↓ and ragged red fibers in the muscle | no data | 400mg/day, response not described | 18 years of age | Young-Mock Lee, University College of Medicine, Korea | https://doi.org/10.26815/acn.2020.00276 |
| **COQ8A [98]** | 1 (1) | R410X (HOM) | no data | 2 y/o (M) | ataxia, dysarthria | no data | no data | no data | 7 years of age | Zhi-Ying Wu, | (Cheng et al., 2021) |

| | | | | | | | | | | | |
|---|---|---|---|---|---|---|---|---|---|---|---|
| **COQ8A [99]** | 1 (1) | R277H/ R301W (CH) | no data | 9 y/o (M) | ataxia, dysarthria, cognition impairment | no data | no data | no data | 11 years of age | Zhejiang University, China | |
| **COQ8A [100]** | 1 (1) | R598H/ S616fs (CH) | no data | 14 y/o (M) | ataxia, head and hands shaking | no data | no data | no data | 17 years of age | | |
| **COQ8A [101]** | 1 (1) | S616fs (HOM) | no data | 24 y/o (M) | ataxia, head and hands shaking, dysphagia | no data | no data | no data | 26 years of age | | |
| **COQ8A [102]** | 1 (1) | L320fs (HOM) | no data | 32 y/o (F) | ataxia, dysarthria, cognition impairment | no data | no data | no data | 52 years of age | | |
| **COQ8A [103]** | 2 (1) | c.656-1G>T (HOM) | no data | 20 y/o (F) | ataxia, writer's cramp | blood lactate in normal range | no data | 60mg/day of ubiquinol, initiated at 20 years old, stopped after only 2 months due to incompliance, no response *[NR]* | 45 years of age | Elisabetta Indelicato, University of Innsbruck, Austria | (Amprosi et al., 2021) |
| **COQ8A [104]** | | c.656-1G>T (HOM) | no data | 7 y/o (M) | ataxia, writer's cramp | no data | no data | 60mg/day of ubiquinol, initiated at 25 years old, due to adverse event (frequent headache); switched to 5mg/kg/day of $CoQ_{10}$; no response at 1-year follow-up *[NR]* | 28 years of age | | |
| **COQ8A [105]** | 1 (1) | A339T (HOM) | no data | 14 m/o (F) | hypotonia, developmental delay, ataxia, glaucoma, dysmorphic features | serum creatine kinase↑; other investigations, including urinary organic acids and blood lactate were unremarkable. ragged-red fibers in the muscle. | CII+CIII↓, CS↑ (muscle) | 100mg/day, response not described | 6 years of age | Robert W. Taylor, Newcastle University, UK | (Cotta et al., 2020) |
| **COQ8A [106]** | 1 (1) | Q343_V344delinsHM/ G244_Q284del (CH) | <2% (muscle), normal range (blood white cells) | 2 y/o (F) | speech difficulties, ataxia, tremors, hypotonia, seizure, hypertension, exercise intolerance | blood lactate↑, blood alanine↑, TCA metabolites in urine↑, ragged-red fibers in the muscle. | CII+CIII↓, CS↑ (muscle) | no data | 16 years of age | | |
| **COQ8A [107]** | 2 (1) | R301W/ E446AfsX33 (CH) | no data | 3 y/o (M) | ataxia, tremors, epilepsy, mild intellectual retardation | no data | no data | 15 mg/kg/day for 6 months, no improvement in motor performance (Timed 25-foot walk test, SARA) *[NR]* | 10 years of age | Tommaso Schirinzi, Bambino Gesù Hospital, Italy | (Schirinzi et al., 2019) |
| **COQ8A [108]** | | R301W/ E446AfsX33 (CH) | no data | 3 y/o (M) | ataxia, mild intellectual retardation | no data | no data | | 7 years of age | | |
| **COQ8A [109]** | 1 (1) | G615D/ L197VfsX20 (CH) | no data | 6 y/o (F) | ataxia, tremors | no data | no data | 15 mg/kg/day for 1 year, improvement in Timed 25-foot walk but no significant change in SARA, gait | 8 years of age | | |

| | | | | | | | | | | |
|---|---|---|---|---|---|---|---|---|---|---|
| **COQ8A [110]** | 1 (1) | R301W/ c.589-3C > G (splice) (CH) | no data | 2 y/o (F) | epilepsy, mild intellectual retardation | no data | no data | analysis parameters and 6 min walking test *[NR]* | 13 years of age | | |
| **COQ8A [111]** | 1 (1) | G27C (HOM) | no data | 2 y/o (F) | seizure, developmental regression, hypothyroidism, mitral regurgitation, mitral valve prolapse, cerebellar atrophy, and epilepsia partialis continua | no abnormality in hematological and biochemical laboratory tests | no data | treated with CoQ$_{10}$ after 11 years of age, dosage unknown, no effect on seizure frequency *[NR]* | 11 years of age | Morteza Heidari, Tehran University of Medical Sciences, Iran | (Ashrafi et al., 2022) |
| **COQ8A [112]** | 1 (1) | L609V (HET) | moderate deficiency in fibroblasts and muscle | (F) | ataxia | no data | CII+CIII↓ (muscle) | 30mg/kg/day from 8 years old, a reduction in ICARS after years of treatment *[Obj.]* | 10 years of age | Rafael Artuch, Hospital Sant Joan de Dèu, Spain | (Pineda et al., 2010) |

### S1.9 Primary CoQ$_{10}$ deficiency due to mutations in the *COQ8B/ADCK4* gene (OMIM *615567) [# of patients: 88]

| Gene [Patient ID] | # of patients (# of families) | Mutation | Level of CoQ$_{10}$ (% of control)[1] | Age at onset (sex) if known | Symptoms | Biochemical tests and muscle pathology | RCC enzymes | CoQ$_{10}$ dose and response[2] | Age at last reported exam or death | Corresponding PI | References |
|---|---|---|---|---|---|---|---|---|---|---|---|
| **COQ8B [1]** | 2 (1) | R178W (HOM) | ~ 11% (EBV-transformed lymphoblasts) | 7 y/o | SRNS | no data | no data | no data | kidney transplant at age 10 | Friedhelm Hildebrandt, Boston Children's Hospital, USA | (Ashraf et al., 2013) |
| **COQ8B [2]** | | R178W (HOM) | ~10% (EBV-transformed lymphoblasts) | 13 y/o | SRNS | no data | no data | no data | kidney transplant at age 15 | | |
| **COQ8B [3]** | 1 (1) | W34X/ T319dup (CH) | no data | 10 y/o | SRNS | no data | no data | no data | kidney transplant at age 14 | | |
| **COQ8B [4]** | 2 (1) | F215Lfs X14/R47 7Q (CH) | no data | 13 y/o | SRNS | no data | no data | no data | kidney transplant at age 15 | | |
| **COQ8B [5]** | | F215Lfs X14/R47 7Q (CH) | no data | 12 y/o | SRNS | no data | no data | no data | kidney transplant at age 13 | | |
| **COQ8B [6]** | 3 (1) | D286G/ E483X (CH) | no data | 14 y/o | SRNS | no data | no data | no data | kidney transplant at age 18 | | |
| **COQ8B [7]** | | D286G/ E483X (CH) | no data | 3 y/o | SRNS | no data | no data | no data | 3 years of age | | |
| **COQ8B [8]** | | D286G/ E483X | no data | 9 y/o | SRNS | no data | no data | no data | 9 years of age | | |

| | | | | | | | | | | | |
|---|---|---|---|---|---|---|---|---|---|---|---|
| | | (CH) | | | | | | | | | |
| **COQ8B [9]** | 2 (1) | R320W (HOM) | no data | 12 y/o | SRNS | no data | no data | no data | ESRF at age 17 | | |
| **COQ8B [10]** | | R320W (HOM) | no data | 20 y/o | SRNS | no data | no data | no data | ESRF at age 23 | | |
| **COQ8B [11]** | 2 (1) | R343W (HOM) | no data | 20 y/o | SRNS | no data | no data | no data | ESRF at age 20 | | |
| **COQ8B [12]** | | R343W (HOM) | no data | 18 y/o | SRNS | no data | no data | no data | ESRF at age 19 | | |
| **COQ8B [13]** | 2 (1) | Q452Hfs (HOM) | ~ 27% (fibroblasts) | 16 y/o | SRNS | no data | no data | no data | 16 years of age | | |
| **COQ8B [14]** | | Q452Hfs (HOM) | ~ 27% (fibroblasts) | 21 y/o | SRNS | no data | no data | no data | 21 years of age | | |
| **COQ8B [15]** | 1 (1) | H400Nfs X11 (HOM) | ~ 8% (EBV-transformed lymphoblasts) | < 1 y/o | SRNS | no data | no data | no data | no data | | |
| **COQ8B [16]** | 1 (1) | R178W (HOM) | no data | 30 y/o (F) | NS/FSGS | proteinuria, hypoalbuminemia, uPCR↑ | no data | 20 mg/kg/day, a decrease in uPCR and stabilization of eGFR *[Obj.]* | no data | Toshiki Doi, Hiroshima University Hospital, Japan | (Maeoka et al., 2020) |
| **COQ8B [17]** | 5 (1) | E447Gfs X10 (HOM) | no data | 14 y/o (M) | SRNS/FSGS | no data | no data | no data | ESRF at the age of 17.7 | Beata S. Lipska-Zietkiewicz, Medical University of Gdansk, Poland | (Atmaca et al., 2017; Korkmaz et al., 2016) |
| **COQ8B [18]** | | E447Gfs X10 (HOM) | no data | 7.3 y/o (F) | SRNS/FSGS, epilepsy | no data | no data | no data | ESRF at the age of 12.6 | | |
| **COQ8B [19]** | | E447Gfs X10 (HOM) | no data | 17 y/o (F) | NS | no data | no data | no data | ESRF at the age of 18 | | |
| **COQ8B [20]** | | E447Gfs X10 (HOM) | no data | 27 y/o (F) | NS | no data | no data | no data | ESRF at the age of 31 | | |
| **COQ8B [21]** | | E447Gfs X10 (HOM) | no data | 7 y/o (F) | NS | no data | no data | no data | 7 years of age | | |
| **COQ8B [22]** | 4 (1) | E447Gfs X10 (HOM) | no data | 25.7 y/o (F) | SRNS/FSGS | no data | no data | 20-30mg/kg/day for 3 months, response not described | 37 years of age, ESRF at the age of 35.4 | | |
| **COQ8B [23]** | | E447Gfs X10 (HOM) | no data | 16.7 y/o (M) | NS | no data | no data | not treated | 25.3 years of age, ESRF at the age of 16.7 | | |
| **COQ8B [24]** | | E447Gfs X10 (HOM) | no data | 13.5 y/o (M) | SRNS/FSGS | no data | no data | not treated | 22.3 years of age, ESRF at the age of 16.6 | | |

| | | | | | | | | | | | |
|---|---|---|---|---|---|---|---|---|---|---|---|
| **COQ8B [25]** | | not tested | no data | 22 y/o (M) | NS | no data | no data | no data | ESRF at the age of 22 | | |
| **COQ8B [26]** | 2 (1) | L98R (HOM) | no data | 5.9 y/o (F) | NS/FSGS, primary nocturnal enuresis | no data | no data | no data | 5.9 years of age | | |
| **COQ8B [27]** | | L98R (HOM) | no data | 13.3 y/o (M) | NS/FSGS, primary nocturnal enuresis | no data | no data | no data | ESRF at the age of 14 | | |
| **COQ8B [28]** | 2 (1) | R178W (HOM) | no data | 14.3 y/o (M) | NS/FSGS, hypermetropia, astigmatism | no data | no data | no data | ESRF at the age of 14.3 | | |
| **COQ8B [29]** | | R178W (HOM) | no data | 9.8 y/o (M) | NS/FSGS, hypermetropia, astigmatism | no data | no data | no data | ESRF at the age of 9.8 | | |
| **COQ8B [30]** | 2 (1) | L98R (HOM) | no data | 13.5 y/o (F) | NS/FSGS, lupus-like symptoms | no data | no data | 20-30mg/kg/day for 22 months, response not described | 20.3 years of age | | |
| **COQ8B [31]** | | L98R (HOM) | no data | 27 y/o (F) | NS/FSGS | no data | no data | 20-30mg/kg/day, response not described | 30 years of age | | |
| **COQ8B [32]** | 4 (1) | E447Gfs X10 (HOM) | no data | 14.9 y/o (M) | NS | no data | no data | no data | ESRF at the age of 14.9 | | |
| **COQ8B [33]** | | E447Gfs X10 (HOM) | no data | 13.2 y/o (F) | NS, epilepsy | no data | no data | no data | ESRF at the age of 13.2 | | |
| **COQ8B [34]** | | E447Gfs X10 (HOM) | no data | 18 y/o (M) | NS | no data | no data | no data | ESRF at the age of 18 | | |
| **COQ8B [35]** | | E447Gfs X10 (HOM) | no data | 9 y/o (M) | NS | no data | no data | treated, dosage and response not described | 9 years of age | | |
| **COQ8B [36]** | 2 (1) | D250N (HOM) | no data | 16.9 y/o (M) | SRNS/FSGS | no data | no data | no data | ESRF at the age of 17.4 | | |
| **COQ8B [37]** | | D250N (HOM) | no data | 13.4 y/o (F) | SRNS/FSGS | no data | no data | no data | ESRF at the age of 13.7 | | |
| **COQ8B [38]** | 1 (1) | F215Lfs X14 (HOM) | no data | 15.1 y/o (M) | SRNS/FSGS | no data | no data | no data | ESRF at the age of 15.8 | | |
| **COQ8B [39]** | 1 (1) | H400Nfs X11 (HOM) | no data | 10.8 y/o (M) | SRNS/FSGS | no data | no data | no data | ESRF at the age of 15.9 | | |
| **COQ8B [40]** | 1 (1) | P310L/ A498E (CH) | no data | 5.1 y/o (F) | NS/FSGS, seizure | no data | no data | no data | ESRF at the age of 13.6 | | |
| **COQ8B [41]** | 1 (1) | F215Lfs X14 (HOM) | no data | 14.2 y/o (M) | NS/FSGS, retinitis pigmentosa, hypospadias | no data | no data | no data | ESRF at the age of 13.6 | | |
| **COQ8B [42]** | 1 (1) | E447Gfs X10 (HOM) | no data | 17.6 y/o (M) | SRNS/FSGS | no data | no data | no data | ESRF at the age of 18 | | |
| **COQ8B [43]** | 1 (1) | E81X/ R490C CH) | no data | 11 y/o (M) | steroid-resistant nephrotic-level proteinuria | no data | no data | no data | normal renal function at age of 12 | Friedhelm Hildebrandt, | (Wang et al., 2017a) |

| | | | | | | | | | | | |
|---|---|---|---|---|---|---|---|---|---|---|---|
| **COQ8B [44]** | 1 (1) | R150X/ D250H (CH) | no data | 8 y/o (F) | SRNS/FSGS | no data | no data | no data | ESRF at the age of 11.7 | Boston Children's Hospital, USA | |
| **COQ8B [45]** | 1 (1) | R178W/ D250H (CH) | no data | 9 y/o (F) | SRNS | no data | no data | no data | ESRF at the age of 11 | | |
| **COQ8B [46]** | 1 (1) | S246N (CH) | no data | 8 y/o (F) | SRNS/FSGS | no data | no data | no data | normal renal function at age of 9 | | |
| **COQ8B [47]** | 1 (1) | S246N (CH) | no data | 17 y/o (F) | isolated proteinuria | no data | no data | no data | normal renal function at age of 18 | | |
| **COQ8B [48]** | 1 (1) | D250H (HOM) | no data | 10 days (F) | NS | no data | no data | no data | no data | | |
| **COQ8B [49]** | 1 (1) | D250H (HOM) | no data | 1 y/o (F) | SRNS/FSGS | no data | no data | no data | ESRF at the age of 6 | | |
| **COQ8B [50]** | 1 (1) | D250H/ Q365E (CH) | no data | 6 y/o (F) | proteinuria/FSGS | no data | no data | no data | normal renal function at age of 12 | | |
| **COQ8B [51]** | 2 (1) | P150Q/ N253K (CH) | no data | 8 y/o (F) | SRNS/FSGS | no data | no data | not treated | ESRF at the age of 15 | Hae Il Cheong, Seoul National University Children’s Hospital, South Korea | (Park et al., 2017b) |
| **COQ8B [52]** | | P150Q/ N253K (CH) | no data | 5 y/o (M) | SRNS | no data | no data | not treated | ESRF at the age of 10 | | |
| **COQ8B [53]** | 1 (1) | S246N/ N253K (CH) | no data | 10 y/o (F) | SRNS/FSGS | no data | no data | not treated | ESRF at the age of 13 | | |
| **COQ8B [54]** | 1 (1) | S246N (HOM) | no data | 10 y/o (F) | SRNS/FSGS | no data | no data | not treated | ESRF at the age of 12 | | |
| **COQ8B [55]** | 1 (1) | S246N/ R490C (CH) | no data | 6 y/o (F) | SRNS/FSGS | no data | no data | not treated | ESRF at the age of 10 | | |
| **COQ8B [56]** | 1 (1) | S246N (HOM) | no data | 12 y/o (F) | NS | no data | no data | △complete remission of proteinuria with cyclosporine treatment; after diagnosis of primary CoQ deficiency, started on $CoQ_{10}$ (30mg/kg/day) with simultaneously tapering doses of steroid and cyclosporine, response not described | 13.5 years of age | | |
| **COQ8B [57]** | 1 (1) | D209H/ C306X (CH) | no data | 14 y/o (M) | NS/FSGS | no data | no data | 150mg/day, a very limited reduction in the severity of urine protein/creatine ratio after 3 months of treatment *[NR]* | no data | Zhangxue Hu, West China Hospital, China | (Yang et al., 2018) |

| | | | | | | | | | | | |
|---|---|---|---|---|---|---|---|---|---|---|---|
| **COQ8B [58]** | 1 (1) | D250Y/ A217T (CH) | no data | 5 y/o (M) | SRNS/FSGS | no data | no data | no data | ESRF at the age of 10 | Benedetta Chiodini, Université Libre de Bruxelles, Belgium | (Lolin et al., 2017) |
| **COQ8B [59]** | 4 (1) | H400Qfs X11 (HOM) | no data | 18 y/o (M) | non-nephrotic proteinuria, CKD | no data | no data | not treated | died at the age of 29 | Fatih Ozaltin, Hacettepe University, Turkey | (Atmaca et al., 2017) |
| **COQ8B [60]** | | H400Qfs X11 (HOM) | no data | 12 y/o (M) | NS, CKD, seizure | no data | no data | 20-30mg/kg/day for 13 months, response not described | 13.5 years of age | | |
| **COQ8B [61]** | | H400Qfs X11 (HOM) | no data | 2 y/o (F) | NS | no data | no data | 20-30mg/kg/day for 10 months, a decrease of proteinuria but no change of eGFR | 4.5 years of age | | |
| **COQ8B [62]** | | H400Qfs X11 (HOM) | no data | 7 y/o (M) | non-nephrotic proteinuria | no data | no data | 20-30mg/kg/day for 10 months, a decrease of proteinuria but no change of eGFR | 8.5 years of age | | |
| **COQ8B [63]** | 2 (1) | H400Qfs X11 (HOM) | no data | 13 y/o (F) | non-nephrotic proteinuria, CKD | no data | no data | 20-30mg/kg/day for 17 months, response not described | 22.4 years of age | | |
| **COQ8B [64]** | | H400Qfs X11 (HOM) | no data | 5 y/o (M) | nephrotic syndrome, CKD | no data | no data | 20-30mg/kg/day for 17 months, response not described | 16.5 years of age | | |
| **COQ8B [65]** | 1 (1) | E447Gfs X11 (HOM) | no data | 12 y/o (F) | NS | no data | no data | not treated | died at 14.8 years of age | | |
| **COQ8B [66]** | 5 (1) | H400Qfs X11 (HOM) | no data | 17.7 y/o (F) | NS | no data | no data | not treated | died at 21.1 years of age | | |
| **COQ8B [67]** | | H400Qfs X11 (HOM) | no data | 4.2 y/o (M) | non-nephrotic proteinuria | no data | no data | 20-30mg/kg/day for 12 months, response not described | 18.3 years of age | | |
| **COQ8B [68]** | | H400Qfs X11 (HOM) | no data | 22.6 y/o (F) | NS, CKD | no data | no data | not treated | 26 years of age | | |
| **COQ8B [69]** | | H400Qfs X11 (HOM) | no data | 7.7 y/o (F) | NS | no data | no data | 20-30mg/kg/day for 14 months, response not described | 11 years of age | | |
| **COQ8B [70]** | | H400Qfs X11 (HOM) | no data | 23.7 y/o (F) | non-nephrotic proteinuria | no data | no data | 20-30mg/kg/day for 10 months, a decrease of proteinuria but no change of eGFR | 24.6 years of age | | |
| **COQ8B [71]** | 3 (1) | E447Gfs X11 (HOM) | no data | 12,4 y/o (F) | protéinurie, ESRF, cardiomyopathy | no data | no data | 20-30mg/kg/day for 15 months, response not described | 15.5 years of age | | |

| | | | | | | | | | | | |
|---|---|---|---|---|---|---|---|---|---|---|---|
| **COQ8B [72]** | | E447Gfs X11 (HOM) | no data | 9.6 y/o (F) | NS, CKD | no data | no data | 20-30mg/kg/day for 22 months, response not described | 12.8 years of age | | |
| **COQ8B [73]** | | E447Gfs X11 (HOM) | no data | 20.3 y/o (F) | non-nephrotic proteinuria, ESKD | no data | no data | 20-30mg/kg/day for 10 months, response not described | 25.3 years of age | | |
| **COQ8B [74]** | 1 (1) | R477Q (HOM) | no data | 17.8 y/o (F) | CKD, autism, hypothy/ooidism, intellectual impairment | no data | no data | 20-30mg/kg/day for 11 months, no response *[NR]* | 20.3 years of age | | |
| **COQ8B [75]** | 1 (1) | K98R (HOM) | no data | 9 y/o (F) | non-nephrotic proteinuria | no data | no data | 20-30mg/kg/day for 21 months, a decrease of proteinuria but no change in eGFR | 18.5 years of age | | |
| **COQ8B [76]** | 3 (1) | E447Gfs X10 (HOM) | no data | 16.4 y/o (F) | NS, CKD | no data | no data | 20-30mg/kg/day for 17 months, response not described | 18 years of age | | |
| **COQ8B [77]** | | E447Gfs X10 (HOM) | no data | 6.4 y/o (M) | CKD, seizure | no data | no data | 20-30mg/kg/day for 16 months, response not described | 26.5 years of age | | |
| **COQ8B [78]** | | E447Gfs X10 (HOM) | no data | 24 y/o (M) | non-nephrotic proteinuria | no data | no data | 20-30mg/kg/day for 13 months, a decrease of proteinuria but no change of eGFR | 25.3 years of age | | |
| **COQ8B [79]** | 3 (1) | K98R (HOM) | no data | 9 y/o (M) | NS | no data | no data | 20-30mg/kg/day for 12 months, response not described | 16.3 years of age | | |
| **COQ8B [80]** | | K98R (HOM) | no data | 9.6 y/o (M) | non-nephrotic proteinuria, ESKF, pulmonary hypertension | no data | no data | 20-30mg/kg/day for 12 months, response not described | 11 years of age | | |
| **COQ8B [81]** | | K98R (HOM) | no data | 32.2 y/o (M) | CKD, pulmonary hypertension | no data | no data | 20-30mg/kg/day for 13 months, a decrease of proteinuria but an increase of eGFR | 39 years of age | | |
| **COQ8B [82]** | 1 (1) | D250H/ R178W (CH) | no data | 9 m/o (F) | proteinuria | no data | no data | 15 to 30mg/kg/day, a reduction of urine protein at 1-year follow-up *[Obj.]* | no data | Jianhua Mao, The Second Hospital of Jiaxing, China | (Feng et al., 2017) |
| **COQ8B [83]** | 1 (1) | D209H/ S205N (CH) | no data | 11 y/o (F) | NS/FSGS, proteinuria | no data | no data | 15 to 30mg/kg/day, no response (proteinuria was persistent, and serum creatine and urea nitrogen were increased at 1-year follow up) *[NR]* | no data | | |
| **COQ8B [84]** | 2 (1) | COQ8B (D250H, HOM) NPHS1 (E447K, HOM) | no data | 9 y/o (F) | SRNS/FSGS, dyspnea, weakness, cardiac dysfunction | anemia, proteinuria, blood BUN and creatinine↑, hypo-albuminemia | no data | △Dosage is not described, given with metoprolol tartrate, losartan potassium, and peritoneal dialysis. At a 2-years follow-up, renal dysfunction was persistent but remained | 11 years of age | Huijie Xiao, Peking University, China | (Zhang et al., 2017) |

| | | | | | | | | | | | |
|---|---|---|---|---|---|---|---|---|---|---|---|
| | | | | | | | | stable, while heart function showed no improvement. | | | |
| **COQ8B [85]** | | COQ8B (D250H, HOM) NPHS1 (E447K, HOM) | no data | 2 y/o (M) | SRNS/FSGS | proteinuria, blood BUN↑, hypo-albuminemia | no data | After the genetic diagnosis, prednisone and tacrolimus were withdrawn and $CoQ_{10}$ treatment started. Renal function showed a slight increase at 2-years follow-up *[NR]* | 2.6 years of age | | |
| **COQ8B [86]** | 1 (1) | R91C/ S246N (HOM) | no data | 3 y/o (M) | Isolated (non-nephrotic) proteinuria | proteinuria, normal eGFR | no data | 15 mg/kg/day, a decrease of proteinuria within 4-months follow-up *[Obj.]* | no data | Li Zhang, The First Hospital of Jilin University, China | (Zhai et al., 2020) |
| **COQ8B [87]** | 1 (1) | I346S/ W520X (CH) | no data | 5 y/o (F) | FSGS, proteinuria, rhabdomyolysis | uPCR↑ | no data | 2100/day since the age of 18 years, developed ESRD a year later *[NR]* | ESRD at 19 years of age | Asmaa S. AbuMaziad, University of Arizona, USA | (AbuMaziad et al., 2021) |
| **COQ8B [88]** | 1 (1) | D250N (HOM) | no data | adolescence (F) | nephropathy, kidney failure | serum creatinine↑ | no data | not treated | kidney transplant at the age of 23 | Mohd Fareed, CSIR Indian Institute of Integrative Medicine, India | (Fareed et al., 2021) |

#### S.1.10 Primary $CoQ_{10}$ deficiency-5 (COQ10D5; 614654) due to mutations in the *COQ9* gene [# of patients: 3]

| Gene [Patient ID] | # of patients (# of families) | Mutation | Level of $CoQ_{10}$ (% of control)[1] | Age at onset (sex) if known | Symptoms | Biochemical tests and muscle pathology | RCC enzymes | $CoQ_{10}$ dose and responses[2] | Age at last reported exam or death | Corresponding PI | References |
|---|---|---|---|---|---|---|---|---|---|---|---|
| **COQ9 [1]** | 1 (1) | R244X (HOM) | ~ 15% (muscle) ~ 18% (fibroblasts) | neonatal (M) | renal tubulopathy, ventricular hypertrophy, seizure, cerebellar atrophy, development delay | blood lactate level↑, type IIB fiber atrophy and lipid accumulation in the muscle | CII+CIII↓ (muscle) | initiated at 11.5 months of age at the dose of 60mg/day and increasing to 300mg/day (after 6 days) which was continued until the patient's death, no response *[NR]* | died at the age of 2 years | Shamima Rahman, Great Ormond Street Hospital, UK | (Duncan et al., 2009; Quinzii and Hirano, 2010; Quinzii et al., 2010; Rahman et al., 2001) |
| **COQ9 [2]** | 1 (1) | S127_R202del (HOM) | ~ 11% (fibroblasts) | neonatal (M) | hypotonia, bradycardia, encephalopathy | blood lactate level↑, blood alanine↑ | CII+CIII↓ (skin) | not treated | died at 18 days of life | H Prokisch, TUM, Germany | (Danhauser et al., 2016) |
| **COQ9 [3]** | 1 (1) | G129Vfs X17 (HOM) | no data | 4 m/o (F) | seizure, hypotonia, dysmorphic features, | no data | no data | initiated at 10 months of age at the dose of 5mg/kg/day and increasing to | 9 months of age | Asburce Olgac, University of Health | (Olgac et al., 2020) |

| | | | | | | | | | | |
|---|---|---|---|---|---|---|---|---|---|---|
| | | | | | growth retardation, microcephaly | | | 50mg/kg/day after the genetic diagnosis, no response *[NR]* | | Sciences, Turkey |

y/o: years old; m/o: months old; HOM: homozygous; HET: heterozygous; CH: compound heterozygous; CSF: cerebrospinal fluid; RCC: respiratory chain complex; CI: complex I; CII: complex II; CIII: complex III; CS: citrate synthase; COX: cytochrome c oxidase; CKD: chronic kidney disease; ICARS: The International Cooperative Ataxia Rating Scale; ETC: electron transport chain; NS: nephrotic syndrome; FSGS: focal segmental glomerulosclerosis; eGFR: estimated Glomerular Filtration Rate; ESRF: end-stage renal failure; ERG: electroretinography; SDH: succinate dehydrogenase; SRNS: steroid-resistant nephrotic syndrome; SND: sensorineural deafness; SARA: Scale for the Assessment and Rating of Ataxia; uPCR: urine protein creation ratio; del: deletion; fs: frameshift; dup: duplication; ins: insertion; delins: deletion-insertion.

[1] CoQ levels are shown as reported or as a percentage relative to the mean value of reported normal range; *[Obj.]*: counted as patients with an objective description of the response to $CoQ_{10}$ treatment in **Table 2**, that is where quantitative or semi-quantitative measures were used to describe $CoQ_{10}$ treatment effects; *[Subj.]*: counted as patients with a subjective description of the response to $CoQ_{10}$ treatment in **Table 2**; *[NR]*: counted as non-responders in **Table 2**; $^{\triangle}$ reported being given simultaneously with other modifications.

**Table S2 Cases excluded from the final analysis and reasons for their exclusion.**

| Gene [Patient ID*] | Mutation | Level of $CoQ_{10}$ (% of control)[1] | Age at onset (sex) if known | Symptoms | $CoQ_{10}$ dose and responses | Reason for exclusion | Reference |
|---|---|---|---|---|---|---|---|
| **COQ2 [1]** | R197H/ N228S (CH) | ~ 36% (fibroblasts) <3% (kidney, muscle) | 18 m/o (M) | SRNS | 30mg/kg/day since age 21 months, response not described | lack of information | (Diomedi-Camassei et al., 2007; Quinzii et al., 2010) |
| **COQ2 [19]** | G390A (HOM) | no data | 18 y/o (F) | SRNS/FSGS | treated, response not described | lack of information | (Gigante et al., 2017) |
| **COQ2 [20]** | G390A (HOM) | no data | 16 y/o (F) | SRNS/FSGS | treated, response not described | lack of information | |
| **COQ4 [6]** | P64S (HOM) | ~ 63% (muscle) | 10 m/o (M) | motor deterioration, ataxia, epileptic seizures, swallowing impairment, progressive scoliosis, cognitive deterioration | treated, response not described | lack of information | (Brea-Calvo et al., 2015) |
| **COQ4 [31]** | G124S (HOM) | low (fibroblasts) | 2 m/o (M) | encephalopathy, spasms, seizure, development delay | beginning at 7 years of age, dose not described, response not described | lack of information | (Yu et al., 2019) |
| **COQ6 [7]** | G255R (HOM) | no data | 0.2 y/o | SRNS, SND, bilateral nephrolithiasis | 30mg/kg/day beginning at 2 months of age (together with enalapril), a decrease of proteinuria, SND and severe growth retardation were noted at 10 months of age | co-treatment with other medication, thus impossible to judge $CoQ_{10}$ treatment effectiveness | (Heeringa et al., 2011) |
| **COQ6 [18]** | P261L (HOM) | no data | 0.8 y/o (M) | SRNS | treated, response not described | lack of information | (Gigante et al., 2017) |
| **COQ6 [25]** | A353D (HOM) | no data | 5 y/o (M) | SRNS, SND, optic atrophy | 15mg/kg/day of idebenone beginning at age of 17 years after the onset of optical symptoms, an improvement in the visual acuity after 2 months of treatment. After 13 months of treatment, the optical examination was stable, but the patient did not recover | insufficient information for judging treatment efficiency | (Justine Perrin et al., 2020) |

| | | | | | | | |
|---|---|---|---|---|---|---|---|
| | | | | | normal vision, still exhibiting persistent optic atrophy. After 3 years of treatment, minimal optic atrophy was reported. No change of the deafness status since treatment initiation. [other medications: immunosuppressive treatment] | | |
| **COQ6 [26]** | A353D (HOM) | no data | 4 y/o (M) | SRNS, SND | 10mg/kg/day of idebenone since age 7, after 13 months of treatment, hearing loss was not changed and renal involvement remained stable with only Enalapril, demonstrated by negative proteinuria. | insufficient information for judging treatment efficiency | (Justine Perrin et al., 2020) |
| **COQ8A [10]** | Y514C/ T584del (CH) | ~ 51% (fibroblasts), ~ 46% (muscle) | 5 y/o (M) | cerebellar ataxia, gynecomastia, feet and thumbs in dystonic position | 60 -700 mg/day over 8 years, the patient reported mild subjective improvement, and stabilization of the cerebellar ataxia was observed on examination | insufficient information for judging treatment efficiency | (Lagier-Tourenne et al., 2008; Lamperti et al., 2003; Quinzii et al., 2010) |
| **COQ8A [17]** | R348X (HOM) | <14.5% (muscle) | 6 y/o (F) | seizure, ataxia, cerebellar atrophy, a mild cognitive delay | 10mg/kg/day initiated at the age of 8 years, within 6 months improvement of ataxia was observed, but after 5 years of treatment, MRI showed increased cerebellar atrophy | insufficient information for judging treatment efficiency | (Terracciano et al., 2012) |
| **COQ8A [19]** | T584delAC C/ P502R (CH) | no data | childhood (F) | mild dysfluent speech and clumsiness, cerebellar atrophy, mild dysarthria | treated, dosage and response not described | lack of information | (Blumkin et al., 2014) |
| **COQ8A [26]** | S616LfsX1 14 (HOM) | no data | 14 y/o (M) | cerebellar ataxia, myoclonus, tremors, dysarthric speech | 200mg/day, improvement in speech and fatigue after 3 months of treatment | insufficient information for judging treatment efficiency | (Liu et al., 2014) |
| **COQ8A [41]** | A338V (HOM) | no data | 13 y/o (F) | cerebellar ataxia, muscle weakness, myoclonus, tremor, dysarthria | dosage not described, improved tremors | insufficient information for judging treatment efficiency | (Traschutz et al., 2020) |
| **COQ8A [42]** | V83fs (HOM) | no data | 8 y/o (F) | cerebellar ataxia, tremors | 1250mg/day, improved tremors | insufficient information for judging treatment efficiency | (Traschutz et al., 2020) |
| **COQ8A [43]** | V83fs (HOM) | no data | 16 y/o (M) | cerebellar ataxia, dysarthria, tremors | 1250mg/day, response not described | lack of information | (Traschutz et al., 2020) |
| **COQ8A [44]** | T584del/ A338T (CH) | ~ 15% (muscle) | 6 y/o (F) | ataxia, pan-cerebellar atrophy | 100mg/day, response not described | lack of information | (Traschutz et al., 2020) |
| **COQ8A [45]** | E481X (HOM) | no data | 1 y/o (F) | ataxia, motor retardation, cognitive impairment, tremors | 200mg/day, no initial apparent effect but after stop: fatigue and falls; improvement of muscle weakness with reintroduction of $CoQ_{10}$ | insufficient information for judging treatment efficiency | (Traschutz et al., 2020) |
| **COQ8A [48]** | c.589-3C>G/ G615D (CH) | no data | 2 y/o (F) | ataxia, hypotonia | 10 mg/kg/day, improvement in stability | insufficient information for judging treatment efficiency | (Traschutz et al., 2020) |

| | | | | | | | |
|---|---|---|---|---|---|---|---|
| **COQ8A [50]** | c.589-3C>G/ R301W (CH) | no data | 2 y/o (F) | ataxia, seizure | 10 mg/kg/day, improved balance | insufficient information for judging treatment efficiency | (Traschutz et al., 2020) |
| **COQ8A [55]** | E568X (HOM) | no data | 6 y/o (F) | spastic hypertonia, ataxia | 300mg/day since 5 years old, more energetic, mentally quicker | insufficient information for judging treatment efficiency | (Traschutz et al., 2020) |
| **COQ8A [60]** | H85AfsX42 (HOM) | no data | 3 y/o (M) | ataxia | 400 - 1200 mg/day, response not described | lack of information | (Traschutz et al., 2020) |
| **COQ8A [74]** | G615D (HOM) | no data | childhood (M) | ataxia, dysmetria, seizure | 135mg/day of idebenone for 9 months, response not described | lack of information | (Mignot et al., 2013) |
| **COQ8A [75]** | G615D (HOM) | no data | 7 y/o (F) | ataxia. dysmetria | 135mg/day of idebenone, response not described | lack of information | (Mignot et al., 2013) |
| **COQ8A [91]** | 27.6 kb deletion of 1q42.3 involving exons 1 and 2 (HOM) | ~ 34% (muscle), normal range (fibroblasts) | 13 y/o (F) | ataxia, tremors, hand bradykinesia, subtle and variable speech dysfluency | Tremor improved on trihexyphenidyl/clonazepam combination therapy before ubiquinol supplementation which was initiated at age 19 years. Ubiquinol dosage was not described. After two years of ubiquinol and high-dose vitamin B-complex treatments, tremor was stable, and the patient was able to tandem walk normally. She had marked bradykinesia though. | insufficient information for judging treatment efficiency | (Galosi et al., 2019) |
| **COQ8A [92]** | G615D/ L197VfsX20 (CH) | no data | 7 y/o (F) | tremors, ataxia, dysmetria, difficulty writing and hand clumsiness | 800mg/day initiated at the age of 8.5, clinical stabilization was reported after the treatment | insufficient information for judging treatment efficiency | (Galosi et al., 2019) |
| **COQ8A [94]** | R301W/ E446AfsX33 (CH) | no data | 3 y/o (M) | ataxia, speech difficulties, seizure, tremors, dystonia | 10 mg/kg/day, initiated at age 10, but has been taken only intermittently, response not described | lack of information | (Galosi et al., 2019) |
| **COQ8A [97]** | c.655+1G>A/ A339T (CH) | no data | 3 y/o (F) | exercise intolerance, dysarthria, seizure, stroke-like episodes, ataxia, homonymous hemianopsia, dysarthria | 400mg/day, response not described | lack of information | https://doi.org/10.26815/acn.2020.00276 |
| **COQ8A [105]** | A339T (HOM) | no data | 14 m/o (F) | hypotonia, developmental delay, ataxia, glaucoma, dysmorphic features | 100mg/day, response not described | lack of information | (Cotta et al., 2020) |
| **COQ8B [22]** | E447GfsX10 (HOM) | no data | 25.7 y/o (F) | SRNS/FSGS | 20-30mg/kg/day for 3 months, response not described | lack of information | (Atmaca et al., 2017; Korkmaz et al., 2016) |
| **COQ8B [30]** | L98R (HOM) | no data | 13.5 y/o (F) | NS/FSGS, lupus-like symptoms | 20-30mg/kg/day for 22 months, response not described | lack of information | (Atmaca et al., 2017; Korkmaz et al., 2016) |
| **COQ8B [31]** | L98R (HOM) | no data | 27 y/o (F) | NS/FSGS | 20-30mg/kg/day, response not described | lack of information | (Atmaca et al., 2017; Korkmaz et al., 2016) |

| | | | | | | | |
|---|---|---|---|---|---|---|---|
| **COQ8B [35]** | E447GfsX 10 (HOM) | no data | 9 y/o (M) | NS | treated, dosage and response not described | lack of information | (Atmaca et al., 2017; Korkmaz et al., 2016) |
| **COQ8B [56]** | S246N (HOM) | no data | 12 y/o (F) | NS | complete remission of proteinuria with cyclosporine treatment; after diagnosis of primary CoQ deficiency, started on $CoQ_{10}$ (30mg/kg/day) with simultaneously tapering doses of steroid and cyclosporine, response not described | lack of information | (Park et al., 2017b) |
| **COQ8B [60]** | H400QfsX 11 (HOM) | no data | 12 y/o (M) | NS, CKD, seizure | 20-30mg/kg/day for 13 months, response not described | lack of information | (Atmaca et al., 2017) |
| **COQ8B [61]** | H400QfsX 11 (HOM) | no data | 2 y/o (F) | NS | 20-30mg/kg/day for 10 months, a decrease of proteinuria but no change of eGFR | | (Atmaca et al., 2017) |
| **COQ8B [62]** | H400QfsX 11 (HOM) | no data | 7 y/o (M) | non-nephrotic proteinuria | 20-30mg/kg/day for 10 months, a decrease of proteinuria but no change of eGFR | | (Atmaca et al., 2017) |
| **COQ8B [63]** | H400QfsX 11 (HOM) | no data | 13 y/o (F) | non-nephrotic proteinuria, CKD | 20-30mg/kg/day for 17 months, response not described | lack of information | (Atmaca et al., 2017) |
| **COQ8B [64]** | H400QfsX 11 (HOM) | no data | 5 y/o (M) | nephrotic syndrome, CKD | 20-30mg/kg/day for 17 months, response not described | lack of information | (Atmaca et al., 2017) |
| **COQ8B [67]** | H400QfsX 11 (HOM) | no data | 4.2 y/o (M) | non-nephrotic proteinuria | 20-30mg/kg/day for 12 months, response not described | lack of information | (Atmaca et al., 2017) |
| **COQ8B [69]** | H400QfsX 11 (HOM) | no data | 7.7 y/o (F) | NS | 20-30mg/kg/day for 14 months, response not described | lack of information | (Atmaca et al., 2017) |
| **COQ8B [70]** | H400QfsX 11 (HOM) | no data | 23.7 y/o (F) | non-nephrotic proteinuria | 20-30mg/kg/day for 10 months, a decrease of proteinuria but no change of eGFR | | (Atmaca et al., 2017) |
| **COQ8B [71]** | E447GfsX 11 (HOM) | no data | 12,4 y/o (F) | proteinuria, ESRF, cardiomyopathy | 20-30mg/kg/day for 15 months, response not described | lack of information | (Atmaca et al., 2017) |
| **COQ8B [72]** | E447GfsX 11 (HOM) | no data | 9.6 y/o (F) | NS, CKD | 20-30mg/kg/day for 22 months, response not described | lack of information | (Atmaca et al., 2017) |
| **COQ8B [73]** | E447GfsX 11 (HOM) | no data | 20.3 y/o (F) | non-nephrotic proteinuria, ESKD | 20-30mg/kg/day for 10 months, response not described | lack of information | (Atmaca et al., 2017) |
| **COQ8B [75]** | K98R (HOM) | no data | 9 y/o (F) | non-nephrotic proteinuria | 20-30mg/kg/day for 21 months, a decrease of proteinuria but no change in eGFR | | (Atmaca et al., 2017) |
| **COQ8B [76]** | E447GfsX 10 (HOM) | no data | 16.4 y/o (F) | NS, CKD | 20-30mg/kg/day for 17 months, response not described | lack of information | (Atmaca et al., 2017) |
| **COQ8B [77]** | E447GfsX 10 | no data | 6.4 y/o (M) | CKD, seizure | 20-30mg/kg/day for 16 months, response not described | lack of information | (Atmaca et al., 2017) |

| | | | | | | | |
|---|---|---|---|---|---|---|---|
| | (HOM) | | | | | | |
| **COQ8B [78]** | E447GfsX 10 (HOM) | no data | 24 y/o (M) | non-nephrotic proteinuria | 20-30mg/kg/day for 13 months, a decrease of proteinuria but no change of eGFR | | (Atmaca et al., 2017) |
| **COQ8B [79]** | K98R (HOM) | no data | 9 y/o (M) | NS | 20-30mg/kg/day for 12 months, response not described | lack of information | (Atmaca et al., 2017) |
| **COQ8B [80]** | K98R (HOM) | no data | 9.6 y/o (M) | non-nephrotic proteinuria, ESKF, pulmonary hypertension | 20-30mg/kg/day for 12 months, response not described | lack of information | (Atmaca et al., 2017) |
| **COQ8B [81]** | K98R (HOM) | no data | 32.2 y/o (M) | CKD, pulmonary hypertension | 20-30mg/kg/day for 13 months, a decrease of proteinuria but an increase of eGFR | not possible to judge treatment efficiency | (Atmaca et al., 2017) |
| **COQ8B [84]** | COQ8B (D250H, HOM) NPHS1 (E447K, HOM) | no data | 9 y/o (F) | SRNS/FSGS, dyspnea, weakness, cardiac dysfunction | dosage is not described, given with metoprolol tartrate, losartan potassium, and peritoneal dialysis. At a 2-years follow-up, renal dysfunction was persistent but remained stable, while heart function showed no improvement. | insufficient information for judging treatment efficiency | (Zhang et al., 2017) |

[*] Patient IDs are the same as in **Table S1**. [1] CoQ levels are shown as reported or as a percentage relative to the mean value of reported normal range; y/o: years old; m/o: months old; HOM: homozygous; HET: heterozygous; CH: compound heterozygous; CKD: chronic kidney disease; ICARS: The International Cooperative Ataxia Rating Scale; NS: nephrotic syndrome; FSGS: focal segmental glomerulosclerosis; eGFR: estimated Glomerular Filtration Rate; ESRF: end-stage renal failure; SRNS: steroid-resistant nephrotic syndrome; SND: sensorineural deafness; SARA: Scale for the Assessment and Rating of Ataxia; uPCR: urine protein creation ratio; del: deletion; fs: frameshift; dup: duplication; ins: insertion; delins: deletion-insertion.

**Table S3 Partial effects reported for CoQ10 treatment of primary CoQ deficiency patients.**

| Gene | Total No. of treated patients [a] | Effects not described or uncertain [a] | No. of patients included in the analysis | Responding | | Not responding |
|---|---|---|---|---|---|---|
| | | | | Objective description | Subjective description | |
| *PDSS1* | 0 | - | 0 | - | - | - |
| *PDSS2* | 2 | 0 | 2 | 0 | 0 | 2 |
| *COQ2* | 10 | 3 | 7 | 1 | 0 | 6 |
| *COQ4* | 21 | 2 | 19 | 3 | 2 | 14 |
| *COQ5* | 3 | 0 | 3 | 3 | 0 | 0 |
| *COQ6* | 5[2] | 2[2] | 3 | 1 | 0 | 2 |
| *COQ7* | 3 | 0 | 3 | 0 | 0 | 3 |
| *COQ8A/ADCK3* | 59[2] | 18[2] | 41 | 9 | 2 | 30 |
| *COQ8B/ADCK4* | 33 | 24 | 9 | 3 | 0 | 6 |
| *COQ9* | 2 | 0 | 2 | 0 | 0 | 2 |

Treatment effects established by quantitative or semi-quantitative measures to describe the response to $CoQ_{10}$ treatment were counted as responding with objective description, while descriptions of convincingly positive effects but without relying on a quantitative or semi-quantitative measure were counted as responding with subjective description. "Not responding" include the patients who were reported not to respond to $CoQ_{10}$ treatment or whose responses we consider lacking a convincing demonstration of a response to $CoQ_{10}$ supplementation. [a] The number of patients treated with the CoQ derivative idebenone are indicated as superscripts.

**Table S4 Patient cases classified as not responding to CoQ10 treatment.**

| Gene [Patient ID*] | Mutation | Level of $CoQ_{10}$ (% of control[1]) | Age at onset (sex) if known | Symptoms | $CoQ_{10}$ dose and responses | Note | Reference |
|---|---|---|---|---|---|---|---|
| **PDSS2 [1]** | Q332X/ S382L (CH) | ~ 14% (muscle) ~ 12% (fibroblasts) | 3 m/o (M) | NS, hypotonia, Leigh syndrome, seizure | 50mg/day beginning at age 3 months, no response, died at age of 8 months *[NR]* | infantile patient with multisystem illnesses, no response observed | (Lopez et al., 2006; Quinzii et al., 2008; Salviati et al., 2012) |
| **PDSS2 [4]** | H162R/ c.1042_1148-2816del (CH) | no data | neonatal (M) | NS, encephalomyopathy, hypertrophic cardiomyopathy, deafness, retinitis pigmentosa, global developmental delay | 20mg/kg/day, no response, died at age of 8 months (1 month after admission) *[NR]* | infantile patient with multisystem illnesses, no response observed | (Ivanyi et al., 2018) |
| **COQ2 [3]** | Y297C (HOM) | ~18% (fibroblasts) ~ 37.5% (muscle) | 11 m/o (M) | infantile encephalomyopathy, SRNS/FSGS, hypotonia, optic atrophy, tremors, psychomotor regression | 30 mg/kg/day beginning at age 22months, neurologic picture improved, but no change in renal function *[NR]* | minimal and ambiguous effects | (Diomedi-Camassei et al., 2007; Montini et al., 2008; Quinzii et al., 2006; Quinzii et al., 2008; Salviati et al., 2005) |
| **COQ2 [4]** | Y297C (HOM) | ~ 17% (fibroblasts) | 12 m/o (F) | NS/FSGS without any clinical signs of neurologic involvement. | 30 mg/kg/day, there was no improvement during the first 2 weeks of treatment; an episode of acute renal failure required continuous hemofiltration for 4 days. 20 days after the initiation of the treatment, recovery of renal function and a reduced level of proteinuria was observed. After 50 months of therapy, renal function remains normal, though proteinuria was still present (other medication: diuretics) *[NR]* | minimal and ambiguous effects | (Diomedi-Camassei et al., 2007; Montini et al., 2008; Quinzii et al., 2006; Quinzii et al., 2008; Salviati et al., 2005) |
| **COQ2 [9]** | S109N (HOM) | ~ 11.4% (fibroblasts) | neonatal (M) | peripheral hypertonia, cardiomyopathy, hypertrophic cardiomegaly, nephrotic syndrome | 30mg/kg/day, no response, died at age of 5 months *[NR]* | infantile patient with multisystem illnesses, no response | (Scalais et al., 2013; Ziosi et al., 2017) |
| **COQ2 [21]** | c.288dupC/ R126G (CH) | no data | 25 y/o (M) | diffuse glomerulosclerosis, end-stage nephropathy, retinopathy | 30 mg/kg/day for 6 months, no ERG improvement, but best corrected visual acuity and areas of retinal atrophy on autofluorescence were noted to be stable on treatment *[NR]* | a minimal and ambiguous effect | (Abdelhakim et al., 2020) |
| **COQ2 [22]** | c.288dupC/ R126G (CH) | no data | 21 y/o (M) | mesangial sclerosis, end-stage nephropathy, retinopathy, lymphoma | | | |
| **COQ2 [23]** | c.288dupC/ R126G (CH) | no data | 23 y/o (F) | retinopathy, end-stage nephropathy | | | |
| **COQ4 [7]** | L82Q/ R158Q (CH) | ~ 16% (muscle) | neonatal (F) | seizures, severe lactic and respiratory acidosis, heart failure | 20 mg/kg/day beginning at the first day of life, which resulted in normalization of lactate and improvement in cardiac function. Nevertheless, the patient continued exhibiting intermittent episodes of lactic acidemia and cardiac decompensation until death (other medications: thiamine, riboflavin, | infantile patient, died shortly despite several treatment attempts | (Chung et al., 2015) |

| | | | | | | | |
|---|---|---|---|---|---|---|---|
| | | | | | hydroxocobalamin, biotin), died at 2 months of age *[NR]* | | |
| **COQ4 [12]** | R240C (HOM) | no data | neonatal (F) | poor/absent reflexes, cardiac hypertrophy, left hip dysplasia, hypotonia, episodes of apnea and bradycardia | 15 mg/kg/day beginning at age 1 month, no response [other medications: pyridoxal phosphate, folinic acid, and riboflavin], died at 7 weeks old *[NR]* | infantile patient, died shortly | |
| **COQ4 [14]** | T77I (HOM) | no data | 4 y/o (M) | tremors, dysarthria, seizure, spastic tetraparesis and ataxia | 1000mg/day beginning at age 13, the 6 min walk test was stable over the period of a year *[NR]* | a minimal and ambiguous effect | |
| **COQ4 [15]** | T77I (HOM) | ~ 22% (fibroblasts) | 9 y/o (F) | seizure, dysarthria, spastic tetraparesis, ataxia | 1000mg/day beginning at age 11, the 6 min walk test was stable over a year, developed a second stroke-like episode at age 14 *[NR]* | a minimal and ambiguous effect | |
| **COQ4 [17]** | G124S (HOM) | ~ 50% (fibroblasts) | neonatal (F) | motor deterioration, weak responsiveness, dystonia, nystagmus, respiratory distress, seizure | 50 mg/kg/day, improvement in seizure, screaming, and respiratory distress, no improvement in nystagmus, dystonia, psychomotor development, and ambulation *[NR]* | minimal and ambiguous effects | (Bosch et al., 2018) |
| **COQ4 [19]** | G95D/ R102H (CH) | ~ 98% (fibroblasts) | 5 y/o (F) | cognitive impairment, dysmetria, spastic ataxia, seizure | 100mg/kg/day of ubiquinol, no response after 6 months (as assessed by the SARA scale) *[NR]* | no response observed | |
| **COQ4 [23]** | G124S/ c.402+1G>C (CH) | low (fibroblasts) | neonatal (M) | encephalopathy, cardiomyopathy, visual and hearing impairment, respiratory failure, apnea, developmental delay | 40 mg/kg/day beginning at 5 months of age, poor response, died at 8 months of age *[NR]* | infantile patient with multisystem illness, died shortly | |
| **COQ4 [24]** | G124S/ c.402+1G>C (CH) | no data | neonatal (M) | cardiomyopathy, respiratory distress, metabolic acidosis | 15 mg/kg/day, no response [other medication: carnitine], died at 2.5 days of age *[NR]* | infantile patient, died shortly after birth | |
| **COQ4 [25]** | G124S (HOM) | no data | neonatal (F) | cardiomyopathy, seizure, developmental delay | treated, dose not described, cardiac function improved gradually and normalized after 10 days [other medication: intravenous immunoglobulin] *[NR]* | a minimal effect | |
| **COQ4 [26]** | G124S/ c.402+1G>C (CH) | no data | neonatal (F) | seizure, apnea, encephalopathy, cardiomyopathy | started at the age of 4 years and 5 months, dose not described, no response observed after 1 month of treatment *[NR]* | no response observed after 1 month of treatment | (Yu et al., 2019) |
| **COQ4 [27]** | | no data | 2 m/o (F) | seizure, respiratory distress, cardiomegaly | started at 1 year of age, dose not described, no response, passed away 1 month later *[NR]* | infantile patient, died shortly after start of $CoQ_{10}$ treatment | |
| **COQ4 [28]** | W184R/ c.402+1G>C (CH) | low (fibroblasts) | 8 m/o (M) | microcephaly, developmental delay, dystonia, visual impairment, oro-motor dysfunction | dose not described, no response *[NR]* | no response observed | |
| **COQ4 [28]** | G124S (HOM) | low (fibroblasts) | infancy (F) | visual impairment, dystonia, spasticity, developmental delay | since 2 years old, dose not described, no response, died at 3.5 years of age *[NR]* | died while on $CoQ_{10}$ treatment | |
| **COQ4 [30]** | G124V/ G124S (CH) | low (fibroblasts) | infancy (F) | encephalopathy, dystonia, spasticity, developmental delay, visual impairment, seizure | beginning at 9 months of age, dose not described, subjective improvement in response [other medication: levetiracetam] *[NR]* | a minimal effect | |
| **COQ6 [6]** | G255R (HOM) | no data | 0.3 y/o | SRNS, SND, facial dysmorphism | 100mg/day, improvement of SND *[NR]* | a minimal effect | (Heeringa et al., 2011) |

| | | | | | | | |
|---|---|---|---|---|---|---|---|
| **COQ6 [17]** | R360W/ c.804delC (CH) | no data | 2 y/o (F) | steroid-resistant glomerulopathy, poor growth | 30 mg/kg/day, remission of glomerulopathy after 1 month of treatment, growth acceleration after 12 months and a reduction of respiratory airway infections *[NR]* | minimal and ambiguous effects | (Koyun et al., 2019; Stanczyk et al., 2018) |
| **COQ7 [1]** | V141E (HOM) | ~ 10% (fibroblasts, muscle) | neonatal (M) | muscular hypotonia, developmental retardation, learning disabilities, hearing impairment, visual dysfunction, not able to sit and walk independently | initially treated with idebenone, switched to $COQ_{10}$ after the diagnosis of a primary $CoQ_{10}$ deficiency (around age of 10 years), dosage unknown, stalling the regression and significantly reducing the pain were noted *[NR]* | minimal effects | (Freyer et al., 2015) |
| **COQ7 [2]** | L111P (HOM) | ~ 70% (fibroblasts) | 14 m/o (F) | spasticity, muscle wasting, inability to walk without support | 22.8 mg/kg/day, no response after 3 months of treatment *[NR]* | no response observed | (Wang et al., 2017b) |
| **COQ7 [3]** | K200IfsX56/ R107W (CH) | ~ 12% (fibroblasts) | neonatal (M) | cardiomyopathy, growth retardation, hypotonia, ptosis, visual impairment, hearing impairment, muscle weakness, infantile spasms | beginning at 2 months of age, and the dose was increased to 20 mg/kg/day at 12 months of life, the patient died around the same time *[NR]* | infantile patient with multisystem illness, no response | (Kwong et al, 2019) |
| **COQ8A [1]** | R213W/ G272V (CH) | ~ 29% (muscle) | 18 m/o (F) | hypotonia, *talus valgus*, developmental delay, seizure, ataxia, epilepsia partialis continua | 20 mg/kg/day (350mg/day) for 8 years, no response *[NR]* | no response observed | (Mignot et al., 2013; Mollet et al., 2008) |
| **COQ8A [2]** | R213W/ G272V (CH) | no data | 2 y/o (F) | hypotonia, seizure, ataxia, developmental delay | 350mg/day for 13 months, no response *[NR]* | no response observed | |
| **COQ8A [3]** | E551K (HOM) | ~ 8% (muscle), normal range (fibroblasts) | 18 m/o (M) | cerebella ataxia, strabismus, muscle weakness, trunk hypotonia, tonic seizure | 5mg/kg/day from age 3 years, 10mg/kg/day from age 4 to 7, no response; followed by 10mg/kg/day of idebenone for 7 months which worsened the patient's conditions *[NR]* | no response observed | (Mollet et al., 2008) |
| **COQ8A [21]** | R271C/ A304T (CH) | normal range (muscle) | 15 y/o (F) | cerebellar ataxia, tremors | 300 mg/day, no response after 6 months *[NR]* | no response observed | (Horvath et al., 2012) |
| **COQ8A [22]** | A304V (HOM) | ~ 8% (muscle) | 27 y/o (F) | cerebellar ataxia, upper-limb myoclonus, seizure, dysmetria, cataract | 300 mg/day, no response after 6 months *[NR]* | no response observed | |
| **COQ8A [23]** | R299W (HOM) | no data | 1 y/o (F) | cerebellar ataxia, seizure, mental retardation, unable to walk by 12 years | 200 mg/day, no response within 2 months *[NR]* | no response observed | |
| **COQ8A [24]** | Y429C/? | ~ 22% (muscle) | 1.5-2 y/o (F) | ataxia, muscle weakness, cognitive impairment, horizontal nystagmus, bilateral dysmetria, tremors | 200 mg/day, no response within 2 months *[NR]* | no response observed | |
| **COQ8A [29]** | D305Y (HOM) | low (muscle) | 5 y/o (M) | developmental delay, intellectual disability, ataxia, isolated pan-cerebellar features including head titubation, dysmetria, dysdiadochokinesia | 800mg/day, inconsistent use for 2 years, no response *[NR]* | no response observed | |
| **COQ8A [38]** | A339T/ Y361 (CH) | no data | 42 y/o (M) | cerebellar ataxia, stroke-like episode, muscle weakness, hearing loss | dosage not described, no response *[NR]* | no response observed | |
| **COQ8A [39]** | A337T (HOM) | no data | 6 y/o (M) | cerebellar ataxia, dystonia, tremor, | 600mg/day, no response *[NR]* | no response observed | |
| **COQ8A** | R301W/ | low | 3 y/o | ataxia | 10 mg/kg/day, no response *[NR]* | no response observed | |

| | | | | | | | |
|---|---|---|---|---|---|---|---|
| **[51]** | E446AfsX33 (CH) | (muscle) | (M) | | | | |
| **COQ8A [52]** | R301W/ E446AfsTer33 (CH) | no data | 2 y/o (M) | ataxia, developmental retardation | 10 mg/kg/day, no response *[NR]* | no response observed | |
| **COQ8A [53]** | R348X (HOM) | low (muscle) | 10 y/o (F) | epilepsy, ataxia | 600mg/day, no response *[NR]* | no response observed | |
| **COQ8A [54]** | R301W (HOM) | low (muscle) | 8 y/o (F) | ataxia, seizure, cardiomyopathy | 400mg/day, no response *[NR]* | no response observed | |
| **COQ8A [78]** | R299W/ R410X (CH) | no data | 4 y/o (F) | ataxia, dysmetria, seizure | 300mg/day for1 month, withdrawn, reversible side effect of treatment (anorexia) *[NR]* | no response observed | |
| **COQ8A [79]** | R299W/ R410X (CH) | no data | 4 y/o (M) | ataxia, dysmetria, seizure | 300mg/day for1 month, withdrawn, reversible side effect of treatment (diarrhea) *[NR]* | no response observed | |
| **COQ8A [80]** | R271C (HOM) | low (plasma) | 1.5 y/o (F) | ataxia, seizure, dystonia, chorea, dysmetria, myoclonus, spasticity | 30 mg/kg/day for 3 years, no response *[NR]* | no response observed | |
| **COQ8A [81]** | L197VfsX20 (HOM) | no data | 19 y/o (F) | ataxia. dysmetria | 1200mg/day for 1 year no response *[NR]* | no response observed | |
| **COQ8A [82]** | L197VfsX20 (HOM) | no data | 19 y/o (F) | ataxia. dysmetria, seizure | 1200mg/day for 1 year, no response *[NR]* | no response observed | |
| **COQ8A [83]** | Q360_Y361ins X (HOM) | no data | 2 y/o (F) | ataxia. Dysmetria, tremors | 800mg/day for 1 year, no response *[NR]* | no response observed | |
| **COQ8A [84]** | R299W (HOM) | ~ 10-24% (muscle) | 7 y/o (F) | ataxia, seizure, tremor | 900mg/day for 6 months, no response *[NR]* | no response observed | (Hikmat et al., 2016) |
| **COQ8A [87]** | R299W (HOM) | no data | 2 y/o (F) | ataxia, epilepsy, seizure, feeding difficulties | 1000mg/day of deoxyubiquinone (probably ubiquinol) since age of 18, no response *[NR]* | no response observed | |
| **COQ8A [96]** | L277P/ c.1506+1G>A (CH) | normal range (plasma) | childhood (F) | ataxia | 20 mg/kg/day, minimal improvement in an ataxia assessment score at 1-year follow-up *[NR]* | minimal effects | |
| **COQ8A [103]** | c.656-1G>T (HOM) | no data | 20 y/o (F) | ataxia, writer's cramp | 60mg/day of ubiquinol, initiated at 20 years old, stopped after only 2 months due to incompliance, no response *[NR]* | no response observed | (Amprosi et al., 2021) |
| **COQ8A [104]** | c.656-1G>T (HOM) | no data | 7 y/o (M) | ataxia, writer's cramp | 60mg/day of ubiquinol, initiated at 25 years old, due to adverse event (frequent headache); switched to 5mg/kg/day of $CoQ_{10}$; no response at 1-year follow-up *[NR]* | no response observed | |
| **COQ8A [107]** | R301W/ E446AfsX33 (CH) | no data | 3 y/o (M) | ataxia, tremors, epilepsy, mild intellectual retardation | 15 mg/kg/day for 6 months, no improvement in motor performance (Timed 25-foot walk test, SARA) | no response observed | (Schirinzi et al., 2019) |
| **COQ8A [108]** | R301W/ E446AfsX33 (CH) | no data | 3 y/o (M) | ataxia, mild intellectual retardation | 15 mg/kg/day for 6 months, no improvement in motor performance (Timed 25-foot walk test, SARA) | no response observed | |
| **COQ8A [109]** | G615D/ L197VfsX20 (CH) | no data | 6 y/o (F) | ataxia, tremors | 15 mg/kg/day for 1 year, improvement in Timed 25-foot walk but no significant change in SARA, gait analysis parameters and 6 min walking test | minimal and ambiguous effects | |

| | | | | | | | |
|---|---|---|---|---|---|---|---|
| **COQ8A [110]** | R301W/ c.589-3C > G (splice) (CH) | no data | 2 y/o (F) | epilepsy, mild intellectual retardation | 15 mg/kg/day for 1 year, improvement in Timed 25-foot walk but no significant change in SARA, gait analysis parameters and 6 min walking test | minimal and ambiguous effects | |
| **COQ8A [111]** | G27C (HOM) | no data | 2 y/o (F) | seizure, developmental regression, hypothyroidism, mitral regurgitation, mitral valve prolapse, cerebellar atrophy, and epilepsia partialis continua | treated with $CoQ_{10}$ after 11 years of age, dosage unknown, no effect on seizure frequency *[NR]* | no response observed | (Ashrafi et al., 2022) |
| **COQ8B [57]** | D209H/ C306X (CH) | no data | 14 y/o (M) | NS/FSGS | 150mg/day, a very limited reduction in the severity of urine protein/creatine ratio after 3 months of treatment *[NR]* | a minimal effect | (Yang et al., 2018) |
| **COQ8B [74]** | R477Q (HOM) | no data | 17.8 y/o (F) | CKD, autism, hypothyroidism, intellectual impairment | 20-30mg/kg/day for 11 months, no response *[NR]* | no response observed | (Atmaca et al., 2017) |
| **COQ8B [83]** | D209H/ S205N (CH) | no data | 11 y/o (F) | NS/FSGS, proteinuria | 15 to 30mg/kg/day, proteinuria was persistent, and serum creatine and urea nitrogen were increased at 1-year follow up *[NR]* | no response observed | (Feng et al., 2017) |
| **COQ8B [84]** | COQ8B (D250H, HOM) NPHS1 (E447K, HOM) | no data | 9 y/o (F) | SRNS/FSGS, dyspnea, weakness, cardiac dysfunction | dosage is not described, given with metoprolol tartrate, losartan potassium, and peritoneal dialysis. At a 2-years follow-up, renal dysfunction was persistent but remained stable, while heart function showed no improvement *[NR]* | no response observed | (Zhang et al., 2017) |
| **COQ8B [85]** | COQ8B (D250H, HOM) NPHS1 (E447K, HOM) | no data | 2 y/o (M) | SRNS/FSGS | After the genetic diagnosis, prednisone and tacrolimus were withdrawn and $CoQ_{10}$ treatment started. Renal function showed a slight increase at 2-years follow-up *[NR]* | a minimal effect | |
| **COQ8B [87]** | I346S/ W520X (CH) | no data | 5 y/o (F) | FSGS, proteinuria, rhabdomyolysis | 2100mg/day since the age of 18 years, developed ESRD a year later *[NR]* | no effect on disease progression | (AbuMaziad et al., 2021) |
| **COQ9 [1]** | R244X (HOM) | ~ 15% (muscle) ~ 18% (fibroblasts) | neonatal (M) | renal tubulopathy, ventricular hypertrophy, seizure, cerebellar atrophy, development delay | initiated at 11.5 months of age at the dose of 60mg/day and increasing to 300mg/day (after 6 days) which was continued until the patient's death, no response *[NR]* | no response observed | (Duncan et al., 2009; Quinzii and Hirano, 2010; Quinzii et al., 2010; Rahman et al., 2001) |
| **COQ9 [3]** | G129VfsX17 (HOM) | no data | 4 m/o (F) | seizure, hypotonia, dysmorphic features, growth retardation, microcephaly | initiated at 10 months of age at the dose of 5mg/kg/day and increasing to 50mg/kg/day after the genetic diagnosis, no response *[NR]* | no response observed | (Olgac et al., 2020) |

[*] Patient IDs are the same as in **Table S1**. [1] CoQ levels are shown as reported or as a percentage relative to the mean value of reported normal range; y/o: years old; m/o: months old; HOM: homozygous; HET: heterozygous; CH: compound heterozygous; CSF: cerebrospinal fluid; RCC: respiratory chain complex; CI: complex I; CII: complex II; CIII: complex III; CS: citrate synthase; COX: cytochrome c oxidase; CKD: chronic kidney disease; ICARS: The International Cooperative Ataxia Rating Scale; ETC: electron transport chain; NS: nephrotic syndrome; FSGS: focal segmental glomerulosclerosis; eGFR: estimated Glomerular Filtration Rate; ESRF: end-stage renal failure; ERG: electroretinography; SDH: succinate dehydrogenase; SRNS: steroid-resistant nephrotic syndrome; SND: sensorineural

deafness; SARA: Scale for the Assessment and Rating of Ataxia; uPCR: urine protein creation ratio; del: deletion; fs: frameshift; dup: duplication; ins: insertion; delins: deletion-insertion.

**Table S5 Cases with positive outcomes following $CoQ_{10}$ treatment, classified as responding.**

| Gene [Patient ID*] | Mutation | Level of $CoQ_{10}$ (% of control)[1] | Age at onset (sex) if known | Symptoms | $CoQ_{10}$ dose and responses | Category of description of $CoQ_{10}$ treatment effects | Reference |
|---|---|---|---|---|---|---|---|
| **COQ2 [25]** | Y353C/ T325A (CH) | no data | 7 m/o (F) | SRNS | 30 mg/kg/ day beginning at age 11 months, urinary protein decreased with the increasing dose of $CoQ_{10}$, and an increase of serum albumin, now on the dosage of 600mg/day *[Obj.]* | objective, as a decrease of proteinuria was reported | (Li et al., 2021) |
| **COQ4 [1]** | mono-allelic deletion (CH) | ~ 43% (fibroblasts) | neonatal (M) | dysmorphic features, mental retardation, encephalomyopathy | 30 mg/kg/day, improvement in physical status and social function. Conditions worsened (weakness and diffuse myalgia) after formulation change and dosage reduction to 2mg/kg/day. Remission of symptoms within a week after reverting back to the original dosage. Then switched to 15mg/kg/day of ubiquinol *[Obj.]* | objective, loss of response after treatment interruption and remission after resuming $CoQ_{10}$ treatment | (Salviati et al., 2012) |
| **COQ4 [18]** | P193S/ R240C (CH) | ~ 95% (fibroblasts) | 2.5 y/o (M) | developmental delay, hypotonia, sialorrhea, spasticity, ataxia | 30 mg/kg/day of ubiquinol, improvement in neuromuscular symptoms after 2 months, further improvement of motor skills in the following months, but speech delay and cognitive impairment persisted *[Subj.]* | subjective, as improvement of more than one symptom was reported | (Mero et al., 2021) |
| **COQ4 [20]** | G55V (CH) | normal range (blood) | 8 y/o (M) | ataxia, spasticity, epilepsy, cognitive deterioration, dysarthria, dysmetria and dysdiadochokinesia | 2000 mg/day, improvement of SARA score, dysarthria is persistent *[obj.]* | objective, as improvement of SARA score was reported | (Caglayan et al., 2019) |
| **COQ4 [21]** | | normal range (blood) | 8 y/o (F) | dysarthria, spastic ataxia, epilepsy, cognitive deterioration, dysmetria, dysdiadochokinesia | Treated, dose not described, improvement of SARA score, gait difficulty and dysarthria are persistent *[obj.]* | objective, as improvement of SARA score was reported | |
| **COQ4 [32]** | G124S (HOM) | no data | 2 m/o (F) | hypotonia, developmental delay, bilateral cortical blinding, seizure, cardiomyopathy | 30mg/kg/day beginning at 11 months of age, some improvement in seizure control and development *[Subj.]* | subjective, as improvement of more than one symptom was reported | |
| **COQ5 [1]** | biallelic duplication of last 4 exons | ~ 57% (muscle) ~ 50% (leukocytes) | childhood (F) | ataxia, dysarthria, seizures, cognitive disability, behavioral problems, epilepsy, myoclonus, dysarthric cerebellar speech, dysmetria, mild tremors and mild lower limb spasticity | dose not described, improvement of ICARS scoring after 3 months, the patient appeared to have a quicker response rate during conversation and better alertness *[Obj.]* | objective, as improvement of SARA score was reported | (Malicdan et al., 2018) |
| **COQ5 [2]** | | ~ 66% (leukocytes) | childhood (F) | mild static gait ataxia, mild dysarthria, mild dysmetria and oculomotor apraxia, and horizontal nystagmus | dose not described, improvement of ICARS scoring after 3 months, the patient appeared to have a quicker response rate during conversation and better alertness *[Obj.]* | objective, as improvement of ICAR score was reported | |

| | | | | | | | |
|---|---|---|---|---|---|---|---|
| **COQ5 [3]** | | ~ 60% (leukocytes) | childhood (F) | mild motor delay, mild learning difficulties, mild cerebellar ataxia, mild cerebellar dysarthria and horizontal nystagmus | dose not described, improvement of ICARS scoring after 3 months, the patient appeared to have a quicker response rate during conversation and better alertness *[Obj.]* | objective, as improvement of ICAR score was reported | |
| **COQ6 [9]** | A353D (HOM) | no data | 2.5 y/o | SRNS, SND | beginning at age 5.5 years, dose not described, decrease of proteinuria but no hearing improvement, reoccurrence of proteinuria after temporary cessation of $CoQ_{10}$ treatment and it decreased again after the treatment resumed *[Obj.]* | objective, loss of response after treatment interruption and remission after resuming $CoQ_{10}$ treatment | |
| **COQ8A [4]** | G272D/ Q605GfsX125 (CH) | < 5% (muscle), normal range (fibroblasts) | 3 y/o (F) | exercise intolerance, muscle weakness, cerebellar syndromes, seizure | 6 mg/kg/day (750mg/day) of $CoQ_{10}$ and L-carnitine were initiated at age 5, improved exercise tolerance and fewer vomiting episodes were noted after 3 months of therapy. $CoQ_{10}$ was replaced with idebenone (5mg/kg/day) at the age of 9 years, and within the following 4 months, severe exercise intolerance reappeared with numerous episodes of vomiting. Reverting to $CoQ_{10}$ treatment resulted in returns to the previous clinical status within 3 months. *[Obj.]* | objective, loss of response after treatment interruption and remission after resuming $CoQ_{10}$ treatment | (Aure et al., 2004; Mignot et al., 2013; Mollet et al., 2008) |
| **COQ8A [18]** | T584delACC/ P502R (CH) | no data | 2 y/o (F) | cerebellar ataxia, dysarthria, nystagmus, cognitive decline, psychiatric disorder | 20mg/kg/day initiated at age 5, partial improvement in motor skills, balance, and strength; after 6 years, treatment was discontinued, and the patient's condition deteriorated. *[Obj.]* | objective, as improvement in more than symptom was reported and the patient's condition worsened after stopping $CoQ_{10}$ treatment | (Blumkin et al., 2014) |
| **COQ8A [20]** | S616LfsX114/ R301Q (CH) | ~ 45% (plasma) | 9 y/o (M) | exercise intolerance, cerebellar ataxia, tremors, dysautonomia | 120mg/day, self-reported fatigue and exercise tolerance improved after 2 weeks of therapy. After 2 years of therapy, ataxia and head tremor diminished and SARA total score improved. When the treatment was stopped for a month, the patient's condition deteriorated, rendering him to resume taking $CoQ_{10}$. *[Obj.]* | objective, as improvement of more than one symptom, including SARA score, was reported; and the patient' condition worsened after stopping $CoQ_{10}$ treatment | (Zhang et al., 2020) |
| **COQ8A [25]** | S616LfsX114 (HOM) | ~ 35% (fibroblasts) | 10 y/o (F) | cerebellar ataxia, myoclonus, slurred speech, wheelchair-dependent by 30 years of age | 400mg/day, improvement in myoclonic symptoms, speech quality (after 3 months), and ataxia with a reduction in SARA (after 6 months) *[Obj.]* | objective, as improvement of more than one symptom, including SARA score, was reported | (Liu et al., 2014) |
| **COQ8A [27]** | R301W/ c.1399-3_ 1408del (CH) | low (muscle) | 11 y/o (M) | reduced dexterity, dysarthria, hypometric saccades, scanning speech, and dystonic posturing, tremors, ataxia | 800mg/day, a resolution of tremors and improvement of limb and truncal dystonia after 9 months of treatment *[Subj.]* | subjective, as improvement of more than one symptom was reported | (Chang et al., 2018) |

| | | | | | | | |
|---|---|---|---|---|---|---|---|
| **COQ8A [28]** | T584del/ T511M (CH) | low (muscle) | 10 y/o (F) | ataxia, tremors, dysarthria, appendicular dysmetria, truncal instability, titubation, wheelchair-dependent by 53 years of age | 800mg/day, improvement of ataxia overall with a reduction in SARA score, able to work independently, after 9 months of therapy. *[Obj.]* | objective, as improvement of more than one symptom, including SARA score, was reported | |
| **COQ8A [76]** | del exons 3-15/ F508S (CH) | no data | 6 y/o (M) | ataxia. dysmetria, myoclonus | 300mg/day for 15 months, improvement in movement disorder and SARA score *[Obj.]* | objective, as improvement of more than one symptom, including SARA score, was reported | |
| **COQ8A [77]** | R299W/ L453RfsX24 (CH) | normal range (fibroblasts) | 15 y/o (M) | ataxia, seizure, myoclonus, dysmetria | 300mg/day for 8 months, improvement in movement disorder *[Subj.]* | subjective, as improvement of more than one symptom was reported | |
| **COQ8A [85]** | R299W/ F578V (CH) | ~ 34-60% (muscle) | 7 y/o (M) | ataxia, seizure, dysmetria, tremors, dysarthria, dysdiadochokinesia | 600mg/day since the age of 33, improvement in balance and coordination (reported by the patient) and a reduction of SARA score *[Obj.]* | objective, as improvement of more than one symptom, including SARA score, was reported | |
| **COQ8A [95]** | L277P/ c.1506+1G>A (CH) | low (muscle) normal range (plasma) | childhood (F) | ataxia, dysmetria, hypotonia | 20 mg/kg/day, improvement in an ataxia assessment score at 1-year follow-up *[Obj.]* | objective, as improvement of an ataxia assessment score was reported | (Jacobsen et al., 2018) |
| **COQ8A [112]** | L609V (HET) | moderate deficiency in fibroblasts and muscle | unknown (F) | ataxia | 30mg/kg/day from 8 years old, a reduction in ICARS after years of treatment *[Obj.]* | objective, as improvement of an ataxia assessment score was reported | (Pineda et al., 2010) |
| **COQ8B [16]** | R178W (HOM) | no data | 30 y/o (F) | NS/FSGS | 20 mg/kg/day, a decrease in uPCR and stabilization of eGFR *[Obj.]* | objective, as improvement of more than one symptom, including uPCR was reported | (Maeoka et al., 2020) |
| **COQ8B [82]** | D250H/ R178W (CH) | no data | 9 m/o (F) | proteinuria | 15 to 30mg/kg/day, a reduction of urine protein at 1-year follow-up *[Obj.]* | objective, as improvement of proteinuria was reported | (Feng et al., 2017) |
| **COQ8B [86]** | R91C/ S246N (HOM) | no data | 3 y/o (M) | Isolated (non-nephrotic) proteinuria | 15 mg/kg/day, a decrease of proteinuria within 4-months follow-up *[Obj.]* | objective, as improvement of proteinuria was reported | (Zhai et al., 2020) |

[*] Patient IDs are the same as in **Table S1**. [1] CoQ levels are shown as reported or as a percentage relative to the mean value of reported normal range; y/o: years old; m/o: months old; HOM: homozygous; HET: heterozygous; CH: compound heterozygous; ICARS: The International Cooperative Ataxia Rating Scale; NS: nephrotic syndrome; FSGS: focal segmental glomerulosclerosis; eGFR: estimated Glomerular Filtration Rate; SRNS: steroid-resistant nephrotic syndrome; SND: sensorineural deafness; SARA: Scale for the Assessment and Rating of Ataxia; uPCR: urine protein creation ratio; del: deletion; fs: frameshift; dup: duplication; ins: insertion; delins: deletion-insertion.

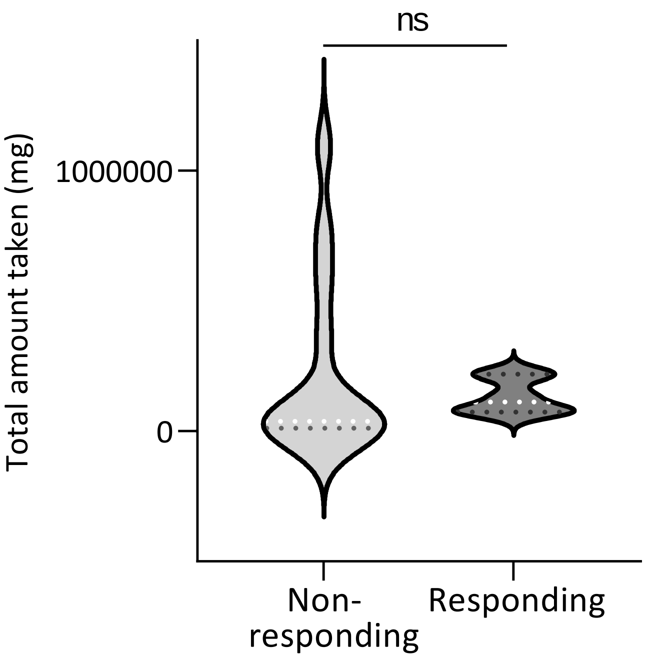

**Fig. S1 The violin plot of total $CoQ_{10}$ amounts taken.** The amounts were calculated as dosage/day x duration. Only the treatments for which $CoQ_{10}$ dosages were reported as mg/day and durations were also reported are included in this analysis. ns: not significant (Student's *t*-test).
